# Assessing ethnic differences in age-standardised net survival of eight common cancer: an English population-based study

**DOI:** 10.64898/2026.08.05.26359765

**Authors:** Tanimola Martins, Bernard Rachet, William Hamilton, Sara Benitez Majano

**Author notes:** Correspondence author’s.

## Abstract

**Background:** We examined ethnic differences in age-standardised net survival (ANS) for eight common cancers diagnosed in England between 2010 and 2019.

**Methods:** Analyses included 247,428 patients aged ≥40 years diagnosed with breast, prostate, lung, colorectal, cervical, ovarian, myeloma, and oesophagogastric cancers. Net survival was estimated at one, three, and five years using the Pohar-Perme estimator and age-standardised with International Cancer Survival Standards weights across four age bands.

**Results:** Compared with White patients, Black patients had higher ANS for lung and prostate cancers at all time points, for myeloma at one year, and for oesophagogastric cancer at one and three years. However, they had lower ANS for breast cancer at three years. Asian patients had higher ANS for lung, prostate, and oesophagogastric cancers at all time points, and for other sites at varying follow-up times. Patients in the Mixed group had higher ANS for most cancers, whereas those in the Other ethnic group generally had lower ANS compared with White patients.

**Conclusions:** Ethnic minority groups in England do not consistently experience poorer cancer survival, with varying patterns observed by cancer site. Universal healthcare access may reduce disparities observed elsewhere, highlighting the importance of context-specific research and public policy.

## Background

In the UK, significant ethnic variations exist in the incidence of several cancers, with myeloma, Hodgkin lymphoma, thyroid, prostate, and certain gastrointestinal cancers all more frequently diagnosed in Asian and Black populations than their White counterparts.[1] What is less clear is whether such differences extend to cancer outcomes, particularly given recent evidence showing statistically significant, albeit modest, ethnic disparities in diagnostic pathways.[2–6] In particular, evidence suggests that the British Asian and Black groups, who generally have lower levels of cancer awareness and less often participate in screening programmes, are more likely to delay seeking medical help.[7–9] These groups also appear more likely to experience longer time to diagnosis[2, 4, 6], which may contribute to their higher odds of advanced-stage diagnoses for certain cancers, including breast, ovarian, uterine, colon, and non-small cell lung cancer.[10] Additionally, they also tend to report poorer healthcare experiences than the White groups.[11, 12] However, limited evidence exists on how these variations impact other key cancer outcomes, such as survival, quality of life, and associated financial burdens on services - factors crucial for healthcare policies and planning.

Studies examining ethnic differences in cancer survival often focus on single cancer types, mainly breast cancer, and are typically restricted to the UK’s three major ethnic groups (Asian, Black and White) excluding the Mixed and Other ethnic groups. For instance, a study of 2,915 women diagnosed with breast cancer before the age of 40 (between 2000 and 2008) found that Black women had lower five-year overall and distant relapse-free survival probabilities compared to White women.[13] Although adjustments were made for tumour characteristics and body mass index, the study did not account for other competing risks of death (e.g., cardiometabolic diseases), which are more common in ethnic minority populations, and may partially explain the observed differences. By contrast, a larger longitudinal study of cases diagnosed between 1998 and 2003 in Southeast England found no significant evidence of ethnic differences in breast cancer-specific survival estimates.[14] Similar findings emerged from an analysis of English cancer registry data, for patients diagnosed between 2002 and 2006, showing little evidence of ethnic differences in one- or three-year age-standardized relative survival estimates.[15]

That study also examined ethnic differences in survival for prostate, lung and colorectal cancer and found better age-standardised survival in Asian and Black patients with lung cancer, with little evidence of ethnic differences in colorectal or prostate cancer.[15] These studies are now relatively dated, with the latter two having significant limitations, particularly regarding missing ethnicity data: around 36% and 25% of ethnicity data was missing in these studies, respectively.[14, 15] Ethnicity data collection within the UK National Health Service (NHS) was inconsistent until the introduction of financial incentives in 2006, which subsequently improved the recording of this variable. In the present study, we use routinely gathered healthcare data to examine ethnic differences in age-standardised net survival of common cancers, focusing on cases diagnosed in England between 2010 and 2019, with follow-up until 2021.

## Methods

### Data sources

This study used data from the Clinical Practice Research Datalink (CPRD-), National Cancer Registration and Analysis Service (NCRAS), Office for National Statistics (ONS) Death Registration, Hospital Episode Statistics (HES), and the Index of Multiple Deprivation (IMD). The CPRD is the world’s largest primary care database, with linkages to the other health and area-based datasets used here.[16, 17] The CPRD-Aurum data used in this study - includes anonymised primary care data from 890 practices in England, with over 28 million patients eligible for linkage to other healthcare databases. The dataset is representative of the English population in terms of geography, deprivation, age, gender, and ethnicity. [16–18] HES contains detailed medical records of all hospital admissions and outpatient appointments, including those at independent sector providers funded by the NHS.[19] The cancer registry data contains records of all tumours diagnosed in England, including information on diagnosis date (i.e., the pathological verification date).[20] The IMD is a composite measure of deprivation based on individuals’ home postcodes, covering several domains of material deprivation, including income, employment, education, skills, health, housing, crime rates, access to services, and living environment.[16, 21]

### Participants

Eligible participants were aged at least 40 years on the date of diagnosis, with an incident cancer recorded in NCRAS between January 2010 and December 2016 or CPRD between January 2017 and December 2019. We excluded male breast cancers due to small numbers and patients diagnosed in an atypical sex (e.g., female prostate cancers). All cases identified from the cancer registry data were followed up until the end of December 2019, while those identified from the CPRD were followed until April 2021 - end of study period.

### Study variables

***Cancer diagnosis:*** Information on cancer diagnoses was obtained from the cancer registry data using the International Classification of Diseases (ICD10) codes for cases diagnosed between 2010 and 2016.[20] For cases diagnosed between 2017 and 2019, data were obtained from the CPRD database using relevant medical codes included in Supplementary File 1. Selected sites included four most common non-skin cancers [lung, breast, prostate, colorectal], three commonly diagnosed in ethnic minority groups [oesophagus, stomach, and myeloma], and two gynaecological cancers [cervical and ovarian]. [1, 15]

***Ethnicity:*** Patients’ ethnicities were initially identified from the CPRD codes, or Hospital Episode Statistics (HES) data where missing in the CPRD, as recommended previously.[5, 22] Utilising the UK census groupings, we extracted and combined all ethnicity records into five major ethnic groups: White (White British, White Irish, Any other White); Asian (Indian, Pakistani, Bangladeshi, Chinese, Other Asian); Black (Black Caribbean, Black African, Other Black); Mixed (White & Black Caribbean, White & Black African, White & Asian, Any other Mixed); and Other. Where there were multiple ethnicity codes for an individual in the CPRD, we assigned the most frequently or most recently recorded codes as in previous studies.[5, 22] Those with missing ethnicity data in the CPRD and HES were excluded.

***Vital status*:** Patients’ vital status (dead or censored/alive) was obtained from the CPRD-linked ONS death registration database, which includes the official date and causes of death.[16, 17] The death registration was available up to June 2019 for cases diagnosed between 2010 and 2016, and up to April 2021 for those diagnosed between 2017 and 2019.

***Other variables:*** Patients’ age, sex (male/female), and socioeconomic status were identified from the CPRD. For age, we assigned everyone a nominal birthday of 1st July, because only birth year was available. Participant’s socioeconomic status was defined using the quintiles of the 2015 and 2019 IMD, for those diagnosed up to 2016 or 2019, respectively. Co-morbidities recorded before diagnosis were identified from the CPRD, using clinical codes relating to 36 long-term conditions described in Cassell *et al*.[23] Patients were categorised into five groups based on the General-outcome weighting Cambridge Multimorbidity Score (CMS),[24] with one group containing those with no included morbidities and the remainder categorised according to quartiles of the CMS score.

***Net survival estimation***: We estimated ethnic differences in age-standardised net survival (ANS) at 1-, 3-, and 5-years post-diagnosis for each cancer type. These time points reflect available follow-up in our cohort: the median follow-up was 3.2 years (maximum 11 years) for cases identified from cancer registry (2010–2016), and 2.3 years (maximum 4.3 years) for those identified from the CPRD (2017– 2019). One-year survival captures short-term outcomes related to diagnostic timeliness and early treatment, while 3- and 5-year survival reflect disease progression, and long-term effectiveness of treatment and follow-up care. Net survival represents survival in a hypothetical setting where death from causes other than the index cancer is deemed impossible, accounting for differences in background mortality (from other causes).[25] As cause-of-death data are unreliable in population-based registries, expected mortality was derived from general population life tables matched on age, sex, calendar period, and deprivation.[25] We used ONS generated lifetables for 2010 - 2021, adjusted for age, sex, deprivation.[26] As these life tables were not corrected for excess mortality associated with COVID-19 (2020-2021), during which mortality rates were higher (Supplementary File 2), we adjusted them to incorporate excess non-cancer mortality for this period. Net survival was estimated by ethnicity and age group using the Complete Approach, with follow-up starting at diagnosis from January 2010. We applied the non-parametric Pohar-Perme estimator, which accounts for informative censoring related to patient characteristics (age, sex or deprivation), implemented in Stata 19 using the stns command.[27] [28] Age standardisation used International Cancer Survival Standards (ICSS) weights across age groups 40-59, 60-69, 70-79, and ≥80 years.[29] For ovarian cancer, three age bands (40-59, 60-69, and ≥70) were used due to small numbers. For cervical cancer, two broader age bands (<60 and ≥60 years) were applied, and ANS was not estimated for the Other ethnic group owing to small sample sizes.

## Results

### Participant characteristics

Our cohort comprised 247,589 patients (162,712 from NCRAS and 86,877 from CPRD). As detailed in Supplementary File S3, we excluded 2,077 patients (n=1,756 from NCRAS, n=321 from the CPRD, plus 84 aged ≥100 years from either database), leaving 247,428 patients for the final analysis. The majority were White (90.3%), with smaller proportions of Black (2.36%), Asian (2.31%), Mixed (2.94%), and Other (2.09%) (Table 1). Around half (50.3%) were males, median age at diagnosis was 70years (range:61-79), with comorbidities recorded in 88% of the patients. At diagnosis, Asian and Black patients were younger and had the highest proportions living in deprived areas. Compared with White patients, the odds of having a recorded comorbidity were higher among Black, Asian, and Mixed groups, but lower among those in the Other ethnic group. The most common cancers were breast (26%), prostate (24%), lung cancer (19%) and colorectal (19%), with marked variations by ethnicity. Prostate cancer was most frequent among Black patients (41%), whereas breast cancer predominated among Asian (37%), Mixed (29%), Other (26%), and White patients (25%) (Table 1).

**Table 1:** Participant characteristics.

|  | White, n(%)<br>223,444 (90.3) | Black, n(%)<br>5,833 (2.36) | Asian, n(%)<br>5,704 (2.31) | Mixed, n(%)<br>7,267 (2.94) | Other, n(%)<br>5,180 (2.09) | All, n(%)<br>247,428 (100) |
| --- | --- | --- | --- | --- | --- | --- |
| **Gender** |  |  |  |  |  |  |
| *Female* | 111,067 (49.7) | 2,323 (39.8) | 3,291 (57.7) | 3,735 (51.4) | 2,585 (49.9) | 123,001 (49.7) |
| *Male* | 112,377 (50.3) | 3,510 (60.2) | 2,413 (42.3) | 3,532 (48.6) | 2,595 (50.1) | 124,427 (50.3) |
| **Age, years** |  |  |  |  |  |  |
| *Median (IQR)* | 71 (62-79) | 62 (54-74) | 64 (54-74) | 68 (59-76) | 68 (59-76) | 70 (61-79) |
| *40-59* | 47,128 (21.1) | 2,553 (43.8) | 2,252 (39.5) | 2,018 (27.8) | 1,478 (28.5) | 55,429 (22.4) |
| *60-69* | 60,955 (27.3) | 1,335 (22.9) | 1,608 (28.2) | 2,092 (28.8) | 1,545 (29.8) | 67,535 (27.3) |
| *70-79* | 65,906 (29.5) | 1,270 (21.8) | 1,219 (21.4) | 2,091 (28.8) | 1,324 (25.6) | 71,810 (29.0) |
| *≥80* | 49,455 (22.1) | 675 (11.6) | 625 (10.9) | 1,066 (14.7) | 833 (16.1) | 52,654 (21.3) |
| **Deprivation (1= most and 5=least deprived)** |  |  |  |  |  |  |
| *1* | 34,978 (15.7) | 2,407 (41.3) | 1,216 (21.3) | 1,365 (18.8) | 692 (13.4) | 40,658 (16.4) |
| *2* | 38,831 (17.4) | 1,861 (31.9) | 1,411 (24.7) | 1,471 (20.2) | 826 (15.9) | 44,400 (17.9) |
| *3* | 44,391 (19.9) | 965 (16.5) | 1,218 (21.4) | 1,390 (19.1) | 1,007 (19.4) | 48,971 (19.8) |
| *4* | 50,905 (22.8) | 376 (6.45) | 957 (16.8) | 1,529 (21.0) | 1,262 (24.4) | 55,029 (22.2) |
| *5* | 54,339 (24.3) | 224 (3.84) | 902 (15.8) | 1,512 (20.8) | 1,393 (26.9) | 58,370 (23.6) |
| **Morbidity score (0=none and 4=highest)** |  |  |  |  |  |  |
| *0* | 25,687 (11.5) | 807 (13.8) | 838 (14.7) | 800 (11.0) | 1,171 (22.6) | 29,303 (11.8) |
| *1* | 50,382 (22.6) | 1,526 (26.2) | 1,289 (22.6) | 1,808 (24.9) | 1,579 (30.5) | 56,584 (22.9) |
| *2* | 53,458 (23.9) | 1,587 (27.2) | 1,479 (25.9) | 1,828 (25.2) | 1,196 (23.1) | 59,548 (24.1) |
| *3* | 57,614 (25.8) | 1,266 (21.7) | 1,481 (25.9) | 1,897 (26.1) | 841 (16.2) | 63,099 (25.5) |
| *4* | 36,303 (16.3) | 647 (11.1) | 617 (10.8) | 934 (12.9) | 393 (7.59) | 38,894 (15.7) |
| **Smoking status** |  |  |  |  |  |  |
| *Never smoker* | 68,176 (30.5) | 2,609 (44.7) | 3,153 (55.3) | 2,453 (33.8) | 1,573 (30.4) | 77,964 (31.5) |
| *Ex-smoker* | 66,139 (29.6) | 870 (14.9) | 616 (10.8) | 2,117 (29.1) | 1,183 (22.8) | 70,925 (28.7) |
| *Current* | 41,786 (18.7) | 1,097 (18.8) | 624 (10.9) | 1,470 (20.2) | 1,048 (20.2) | 46,025 (18.6) |
| *Unknown* | 47,343 (21.2) | 1,257 (21.6) | 1,311 (22.9) | 1,227 (16.9) | 1,376 (26.6) | 52,514 (21.2) |
| **Cancer type** |  |  |  |  |  |  |
| *Breast* | 56,124 (25.1) | 1,395 (23.9) | 2,127 (37.3) | 2,130 (29.3) | 1,337 (25.8) | 63,113 (25.5) |
| *Lung* | 43,378 (19.4) | 554 (9.50) | 730 (12.8) | 1,196 (16.5) | 923 (17.8) | 46,781 (18.9) |
| *Prostate* | 52,683 (23.6) | 2,373 (40.7) | 1,090 (19.1) | 1,867 (25.7) | 1,228 (23.7) | 59,241 (23.9) |
| *Colorectal* | 42,009 (18.8) | 689 (11.8) | 937 (16.4) | 1,215 (16.7) | 1,011 (19.5) | 45,861 (18.5) |
| *OG* | 15,171 (6.79) | 283 (4.85) | 298 (5.22) | 402 (5.53) | 369 (7.12) | 16,523 (6.68) |
| *Cervical* | 1,886 (0.84) | 52 (0.89) | 73 (1.28) | 67 (0.92) | 54 (1.04) | 2,132 (0.86) |
| *Myeloma* | 5,745 (2.57) | 385 (6.60) | 207 (3.63) | 211 (2.90) | 104 (2.01) | 6,652 (2.69) |
| *Ovary* | 6,448 (2.89) | 102 (1.75) | 242 (4.24) | 179 (2.46) | 154 (2.97) | 7,125 (2.88) |
| **Vital status** |  |  |  |  |  |  |
| *Alive* | 124,647 (55.8) | 4,155 (71.2) | 4,109 (72.0) | 4,876 (67.1) | 2,965 (57.2) | 140,752 (56.9) |
| *Dead* | 98,797 (44.2) | 1,678 (28.8) | 1,595 (27.9) | 2,391 (32.9) | 2,215 (42.8) | 106,676 (43.1) |

*IQR = Interquartile Range; OG = Oesophagogastric cancer*

### Ethnic differences in vital status

Overall, 43% of participants (106,676/247,428) died during follow-up, with a higher proportion recorded for those diagnosed in 2010 (61%) compared to those in 2019 (24%). The pattern also varied by ethnicity and cancer type: proportions were generally higher among patients with lung or oesophagogastric cancers, and among White and Other ethnic groups (Supplementary File S4). In multivariable Poisson regression, adjusted for age, sex, deprivation, comorbidity, smoking status, cancer type, and year of diagnosis, overall mortality was lower among Black (adjusted risk ratio= 0.89, 95%CI: 0.85-0.93), Asian (0.85, 95% CI: 0.81-0.89), and Mixed group (0.91, 95%CI: 0.87-0.95) but modestly higher in the Other ethnic group (1.08, 95%CI 1.04-1.13) compared with White patients.

### Ethnic differences in Age-specific net-survival by cancer sites

Across all cancer sites and ethnic groups, net survival declined with increasing follow-up time and advancing age, with the highest survival observed among patients aged 40–59 years and the lowest among those aged ≥80years (supplementary file S5). Stratification by ethnicity revealed substantial heterogeneity by cancer type and length of follow-up. Compared with White patients, Black patients had consistently higher net survival across all time points and age groups for prostate cancer, myeloma, and oesophagogastric cancers. By contrast, findings for breast, cervical, and ovarian cancers were less consistent. In particular, Black women aged 40–79 years with breast cancer, those aged 40–69 years with ovarian cancer, and those aged 50–59 years with cervical cancer all had lower survival than their White counterparts, whereas estimates among older age groups were more variable. Asian patients generally had higher survival across most age groups compared with White patients, although estimates were less consistent for myeloma and ovarian cancers. Patients in the Mixed group also tended to have higher survival estimates, whereas those in the Other ethnic group generally had lower survival than White patients.

### Ethnic differences in Age-standardised net-survival (ANS) by cancer sites

The results of ANS by ethnicity and cancer site are presented in Figures 1a–1h. Across all sites, the highest survival was observed for breast and prostate cancers, with 1-year ANS of 95% for both, declining to 79% and 78% at 5years, respectively. One-year ANS was also high for myeloma (83%), but this declined considerably to 50% by 5years post-diagnosis. Colorectal, cervical, and ovarian cancers all had intermediate survival, with 1-year ANS of 79%, 77%, and 72%, decreasing to 54%, 54%, and 41% at 5 years, respectively. In contrast, lung and oesophagogastric cancers had the lowest survival overall, with 1-year ANS of 42% and 48%, falling sharply to 16% and 19% at 5years.

**Figure.**
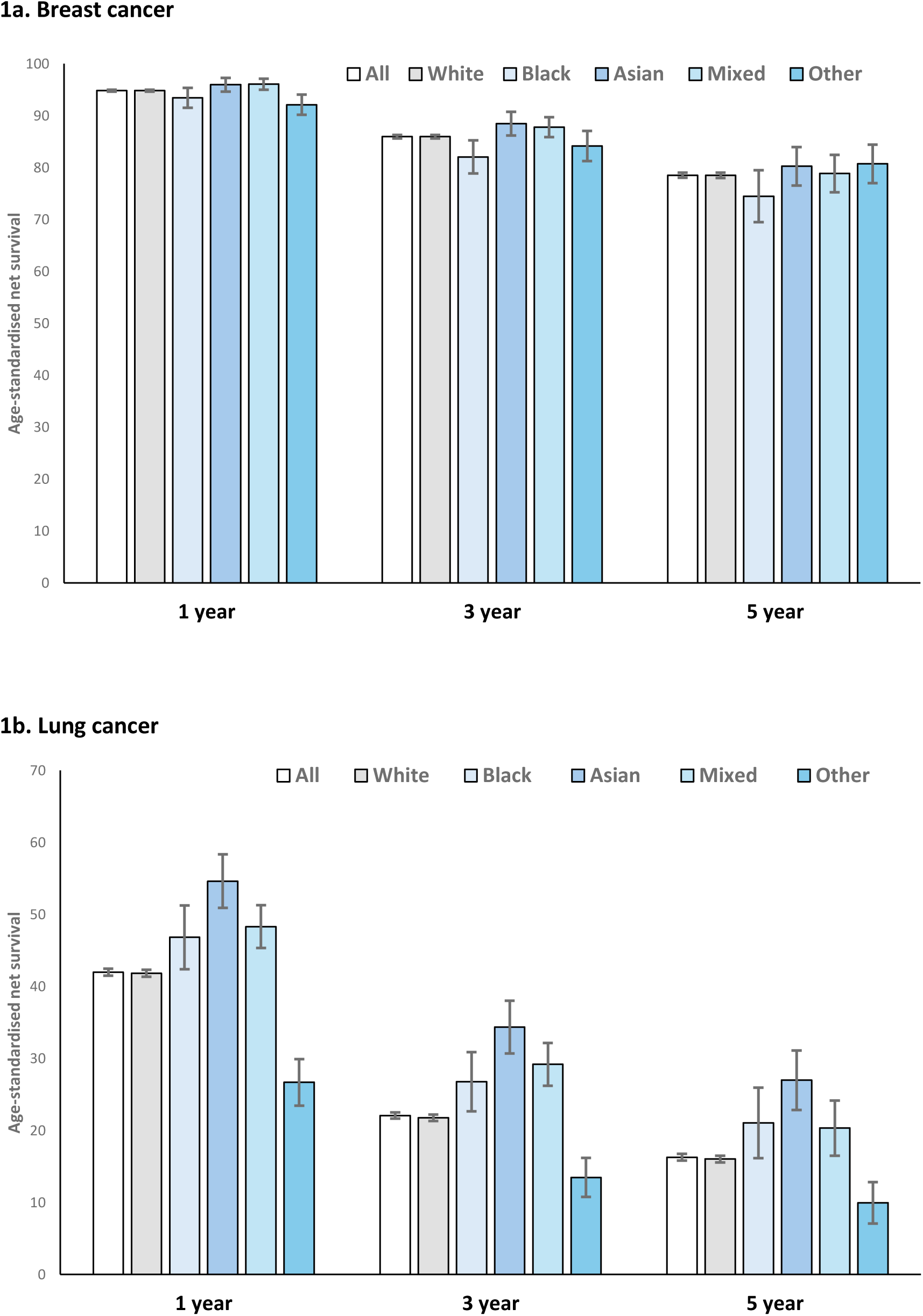
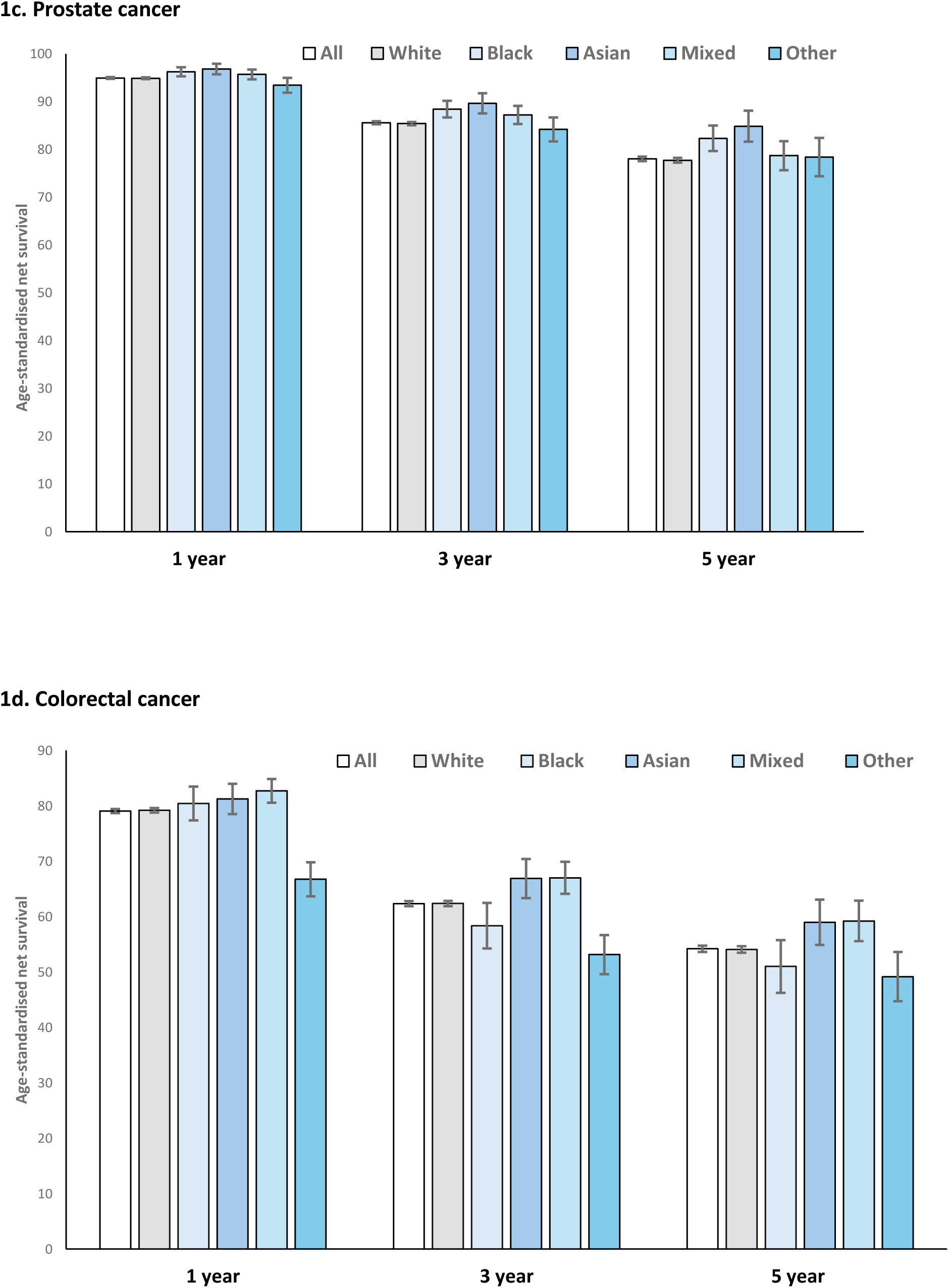
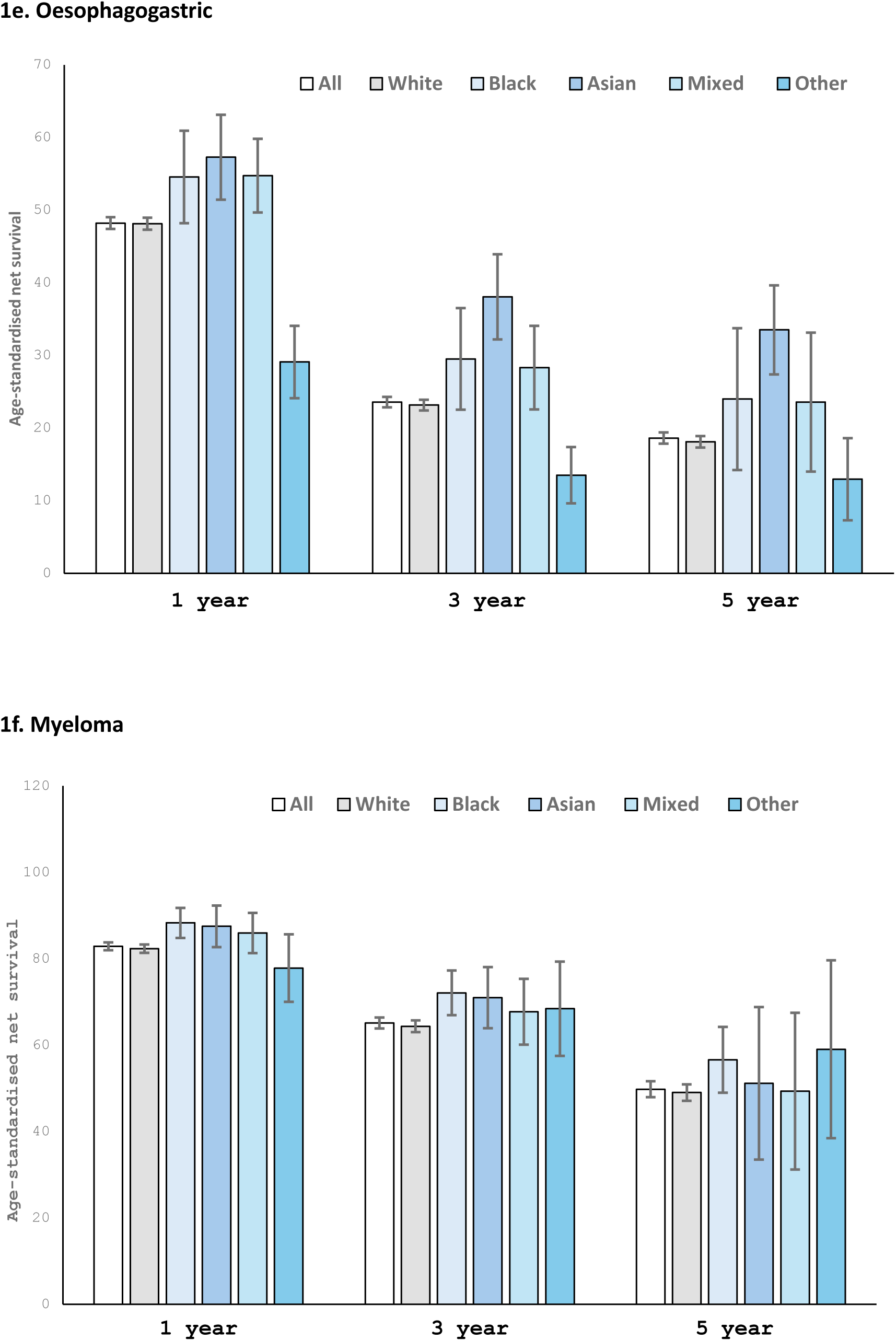
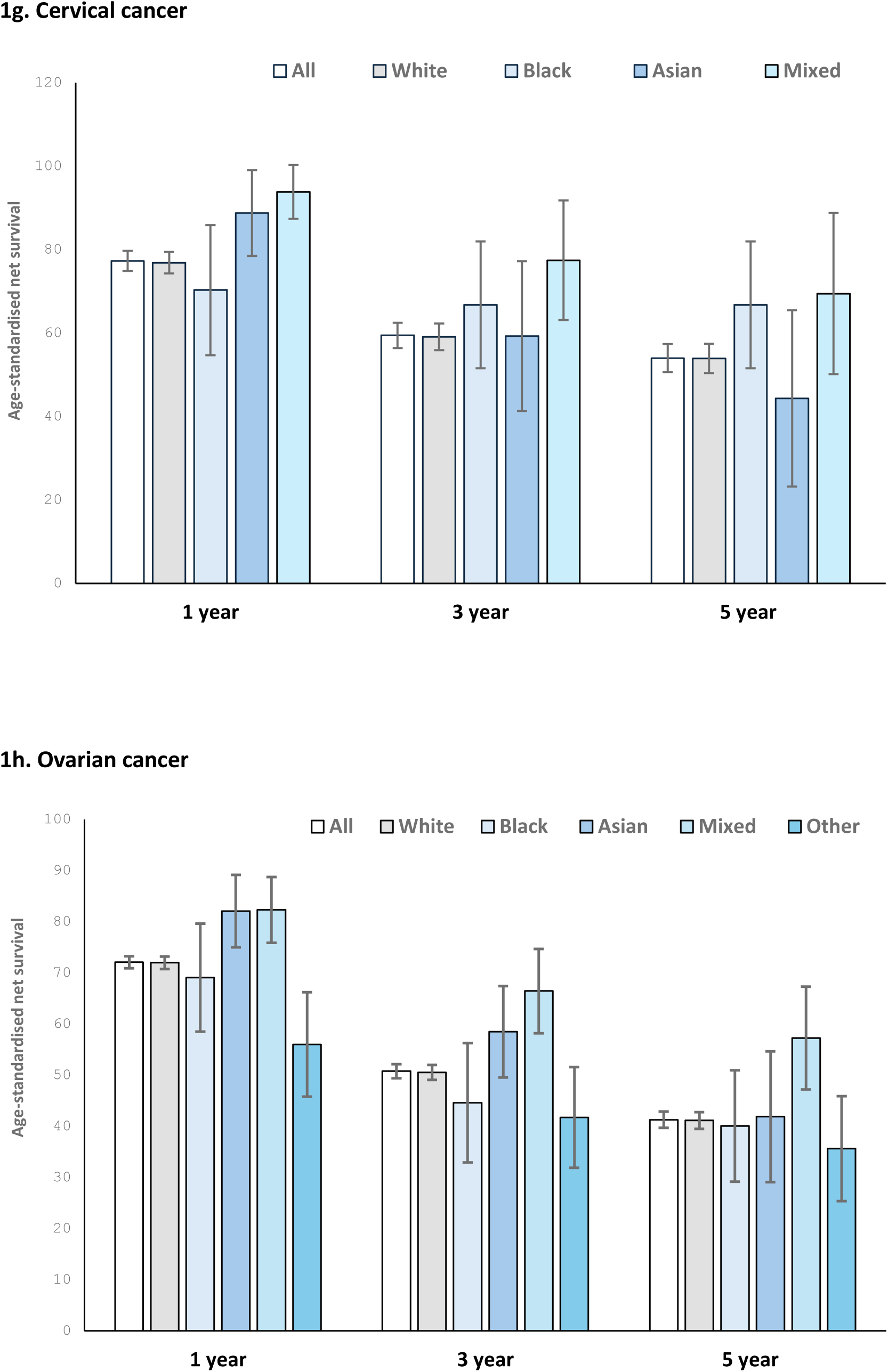
**1a-1h:** Showing ethnic differences in age-standardised Pohar Perme net-survival by cancer type - estimated for patients diagnosed in England between 2017 and 2021 using population lifetables adjusted for age, sex, and deprivation-specific mortality. Estimates are age-standardised; error bars represent 95% confidence intervals.

**Black vs White patients:** Across all time points, ANS was significantly higher among Black than White patients for lung and prostate cancers, with the opposite pattern observed for breast cancer, although the difference was only significant at 3years [82% vs 86%]. Black patients had higher ANS for oesophagogastric cancer at 1year [55% vs 48%] and for myeloma at 1year [88% vs 83%%] and 3years [72% vs 65%]. ANS estimates for colorectal, cervical, and ovarian cancers appears lower among Black compared with White patients, but the differences were not statistically significant.

**Asian vs White patients:** ANS was significantly higher among Asian than White patients for lung, prostate, and oesophagogastric cancers across all time points. Asian patients also had higher ANS for breast cancer patients at 3years [89% vs 86%], for colorectal cancer at 3years [67% vs 62%] and 5years [59% vs 54%], and for myeloma at 1year [88% vs 82%] and 3years [71% vs 64%]. For cervical and ovarian cancers, ANS was higher for Asian patients at 1year, with little evidence of differences at later time points.

**Mixed vs White patients**: ANS was significantly higher among patients in the Mixed group than among White patients for lung, colorectal, and ovarian cancers across all time points. The Mixed group also had higher ANS for breast cancer at 1year [96% vs 95%] and 3years [88% vs 86%], for cervical cancer at 1year [94% vs 77%] and 3years [78% vs 59%], and for oesophagogastric cancer at 1year [55% vs 48%]. There was little evidence of differences in ANS between the Mixed and White groups for myeloma or prostate cancer at any time point.

**Other vs White patients:** Across all time points, ANS was significantly lower among patients in the Other group than White patients for lung, colorectal and oesophagogastric cancers. The Other group also had lower ANS at 1year for breast [92% vs 95%] and ovarian cancer [56% vs 71%], with little evidence of differences in ANS for myeloma and prostate cancers at any time point.

## Discussion

After age adjustment, we observed ethnic differences in ANS by cancer type, although the overall pattern was inconsistent. Compared with White patients, Asian and Black patients generally had higher ANS, particularly for lung, prostate, and oesophagogastric cancers, whereas differences were small or negligible for colorectal, cervical, and ovarian cancers. Patients in the Mixed ethnic group also tended to have higher ANS compared with White patients across several cancer types, although the magnitude of these differences varied. Lower ANS was relatively uncommon in the ethnic minority groups; observed mainly among Black patients with breast cancer and among patients in the Other ethnic group, who tended to have lower ANS across several cancers.

### Interpretation of findings

For breast cancer, our overall 1-year ANS (95%) aligns with previous reports (96%), but our 5-year estimate (76%) was lower than the previously reported 85%.[30] This difference likely reflects the older age distribution of our cohort (≥40 years vs ≥18 in previous work), and adjustments for COVID-19- related excess mortality. Despite higher odds of advanced-stage diagnosis,[10] Asian women in our cohort had higher ANS than White women across all time points, reaching statistical significance at 3years. This pattern may indicate better treatment tolerance or more therapy-responsive tumours.[31] In contrast, Black women had lower ANS than White women at all time points, with a significant difference at 3years. Potential explanations include later–stage presentation, more aggressive tumour subtypes, reduced adherence to adjuvant therapy or suboptimal follow-up care.[31] [32] Our findings of higher short-term ANS among patients in the Mixed group and lower ANS among those in Other ethnic group have not been previously reported. These findings highlight the need for interventions that extend beyond improving screening uptake and early symptomatic diagnosis, to ensuring equitable access to treatment, supporting adherence, and strengthening continuity of care throughout the cancer pathway.

For lung cancer, our overall ANS estimates were similar to previous reports.[30] [15] The higher ANS observed among Asian and Black patients compared with White patients also mirrors earlier findings,[30] [15] [33] although the underlying mechanisms remain unclear, particularly given the higher odds of advanced-stage lung cancer in some Black patients.[10] One potential explanation is the “salmon bias effect,” where seriously ill immigrants return to their country of origin, [33, 34] although this could not be assessed from our data. Differences in age distribution may also contribute: a larger proportion of Black (61%) and Asian patients (51%) were aged ≤60years at diagnosis compared with White patients (39%). Smoking patterns may further explain the survival advantage among Asian patients, who had the highest proportion of never-smokers/ex-smokers (54%) and the lowest proportion of current smokers(26%) (Supplementary File S6 & S7), a factor strongly associated with reduced lung cancer mortality.[35] However, this does not account for the higher ANS observed among Black patients, among whom smoking prevalence was higher (current smokers in Black was 46% vs 38% in White). Further research using more granular ethnicity is needed to better explain these patterns. Our study is the first to report ANS for lung cancer among Mixed and Other ethnic groups.

For prostate cancer, our overall ANS estimates are consistent with previous reports,[30] but there are notable ethnic variations. Asian and Black patients had higher ANS than White patients at all time points, contrasting previous reports showing no survival differences by ethnicity.[15] These patterns contribute additional evidence to ongoing discussions about the potential role of targeted screening for Black men, based on the belief that outcomes are worse in this group. Much of the literature on prostate cancer disparities originates from the US, where the disease burden is higher among Black African Americans. However, emerging evidence from the UK indicates that Black men are less likely to be diagnosed with advanced-stage prostate cancer,[10] suggesting that findings from the US context may not translate directly to the UK healthcare setting. If targeted screening strategies are considered in the UK, then careful evaluation of both potential benefits and harms (e.g., overdiagnosis and overtreatment) will be important, particularly as our results suggest that outcomes are not worse for this group. Our findings for the Mixed and Other ethnic groups are novel.

For colorectal cancer, our overall ANS estimates were similar to previous reports.[30] In contrast to earlier studies,[15] Asian patients in our cohort had higher ANS, particularly at 3- and 5years post-diagnosis, probably reflecting their higher odds of diagnosis via screening and fewer emergency diagnoses compared with White patients.[5] Despite reports of more advanced-stage colon cancer among Black patients,[10] we found no clear differences in ANS between Black and White patients; the estimates were lower for Black patients but not statistically significant. Due to sample size constraints, we did not separate colon and rectal cancers in our analysis. Efforts to improve outcomes for Black patients should focus on increasing screening uptake and minimising diagnostic delays among symptomatic patients. Our findings of consistently higher ANS for the Mixed group and lower ANS for the Other group are novel.

For oesophagogastric cancer, our overall ANS of 48% at 1year, declining to 19% at 5years are consistent with previous estimates for stomach and oesophageal cancers.[30] Asian patients had better ANS than White patients across all time points, with similar patterns observed among Mixed and Black patients, although with statistical significance at 1year only. While the reason for these variations warrants further investigation, the reality is that outcomes for this cancer have remained poor and highlight the urgent need for innovations to improve early detection and more effective treatment. Again, our findings of consistently lower ANS for the Other group have not been previously reported.

For cervical cancer, our overall ANS estimates are slightly lower than previous estimates.[30] Asian patients had higher 1-year but lower 5-year ANS than White patients, although differences at 3 and 5 years were not statistically significant. While Asian patients were as likely as White patients to be diagnosed via screening or emergency presentation, they were more often diagnosed following symptomatic presentation in primary care.[5] It is possible this help-seeking behaviour identifies earlier-stage disease, potentially contributing to better short-term outcomes in Asian patients. However, this advantage did not persist, with lower ANS observed for Asian patients by 5years, though not statistically significant. Our ANS estimates for Black patients appeared lower at 1year, but higher 3- and 5-year compared with White patients, again not statistically significant. These patterns point to the potential influence of other predictors of longer-term survival - such as stage at diagnosis, treatment differences, or access to follow-up care - at play. Improving cervical cancer outcomes must begin with encouraging prevention by vaccination and boosting screening rates, particularly among marginalised ethnic minority populations who are less likely to participate.[36, 37] However, this must be complemented by inclusive early diagnosis initiatives and equitable long-term follow-up care. Our findings of higher ANS at 1 and 3years for the Mixed group are novel.

For ovarian cancer, our overall ANS of 72% at 1year dropping to 41% at 5years are slightly lower than previous estimates (78%-52%).[30] While short-term survival appears high, the steep drop by five years post-diagnosis may point to effectiveness of treatments, particularly for advanced-stage disease, which remains common.[10, 30] Patients in the Mixed group had the highest ANS across all time points, a novel and potentially important finding that must be interpreted cautiously due to the heterogeneity within this group. Asian patients had higher ANS than White patients across all time points, with significant findings at 1year. Black patients had lower ANS across all time points than White patients, although not statistically significant, partly due to small sample sizes. Nevertheless, the trend is concerning and aligns with wider literature pointing to poorer outcomes of ovarian cancer among Black populations, including late-stage diagnosis.[10] Addressing these disparities requires targeted efforts to improve awareness, access to specialist care, and continuity of care in underserved communities.

For myeloma, our overall ANS are consistent with previous estimates.[30] Asian and Black patients had higher ANS across all time points than White patients, although with statistical significance only at 1 and 3years post-diagnosis. There was little evidence of differences in ANS among patients in the Mixed and Other ethnic groups compared with White patients. While these findings are novel and may indicate ethnic variation on the disease subtypes, they should be interpreted cautiously given the small sample sizes in non-White ethnic minority groups.

### Strengths and limitations

The study had a large sample size and used high-quality data obtained from the CPRD and NCRAS databases. Patients’ ethnicity - defined in line with UK national census groupings - were identified from the CPRD and HES, with 99% completeness, allowing more detailed analysis by ethnicity than for previous studies. For instance, this is the first study to examine ANS in the Mixed and Other ethnic groups, though frequently these were too small to allow robust conclusions. However, using combined ethnic categories hides heterogeneity within ethnic subgroups. The alternative would have been to use the 19 ethnic sub-groups in national census, which would have generated many smaller groups, often insufficiently powered. Our analysis was limited to patients aged at least 40 years, recognising that the incidence of certain cancers (e.g., breast and prostate) in the under 40s may be higher in some ethnic minority groups. Nevertheless, the majority of cancer diagnoses across populations occur after age 50, with risk increasing from age 40. Including younger individuals would likely yield small case numbers or introduce more atypical cancers (e.g., more aggressive subtypes), thereby, compounding the within cancer heterogeneity further. Future studies should, however, investigate outcomes among younger cancer populations if they can achieve adequate power. We used the English population lifetable defined by year, age, sex, and deprivation during the study period (with adjustment for COVID- 19-related mortality), as ethnicity-specific life tables were not available. Finally, we only reported ethnic differences in ANS at one to five-years post-diagnosis, whereas 10-year ANS could offer more comprehensive outlook.

## Conclusions

Overall, our findings challenge the assumption that UK ethnic minority groups experience *uniformly poorer outcomes* for common cancers, a pattern frequently reported in US-based research. This perception has been particularly strong with respect to prostate cancer, where worse survival among Black men has been consistently reported in US-based studies. With the exception of breast cancer in Black patients, where survival was slightly worse, we did not observe any other instance in which White patients had significantly or consistently better survival. This highlights the importance of context-specific research when shaping healthcare policies and public health messaging. Structural, cultural, and healthcare system differences between countries such as the UK and the US likely contribute to differing survival patterns across ethnic groups. By leveraging comprehensive national data, our study provides new insights into ethnic differences in cancer survival in the UK. However, limitations such as the absence of ethnicity-specific life tables, small sample sizes in some cancers (e.g., myeloma, cervical and ovarian), and the use of broad ethnic categories highlight areas for future refinement. In particular, although we reported ANS for patients in the Mixed and Other groups, the substantial heterogeneity within these categories necessitates cautious interpretation. Future research should prioritise the development of more granular data, including detailed ethnic classifications and life tables, to enable more accurate and equitable survival estimates. To improve cancer outcomes in the UK, research, public policies and practice should foster inclusivity across all stages of the cancer continuum, ensuring that disparities in diagnosis, treatment, and follow-up care are effectively addressed.

## Authors’ contributions

TM and WH, were involved in all aspects of the study. BR and SB participated in the study design, data analysis, interpretation, preparation and revision of the manuscript. All authors read and approved the final manuscript.

## Ethical approval

The study was approved by the Clinical Practice Research Datalink’s Independent Scientific Advisory Committee (reference number 20_016). Due to the retrospective nature of the study, the need to obtain the informed consent was waived by the Clinical Practice Research Datalink’s Independent Scientific Advisory Committee. All methods were performed in accordance with the relevant guidelines and regulations.

## Data sharing

The data supporting the findings of this study are available from the Clinical Practice Research Datalink (CPRD) subject to licensing restrictions and are therefore not publicly available. Access to the data may be requested directly from CPRD; further information is available from the corresponding author.

## Data Availability

The data supporting the findings of this study are available from the Clinical Practice Research Datalink subject to licensing restrictions and are therefore not publicly available. Access to the data may be requested directly from Clinical Practice Research Datalink; further information is available from the corresponding author.

