## Supplementary files for "Assessing ethnic differences in age-standardised net survival of eight common cancer: an English population-based study"

**S1. CPRD Medical codes and International Classification of Diseases (ICD10 codes) for selected cancers.**

| **CPRD Medical code** | **Description** | **ICD 10** |
| --- | --- | --- |
| 61695 | Malignant neoplasm of cervical oesophagus | C150 |
| 1062 | Malignant neoplasm of oesophagus | C150-C159 |
| 41362 | Malignant neoplasm of thoracic oesophagus | C151 |
| 63470 | Malignant neoplasm of abdominal oesophagus | C152 |
| 50789 | Malignant neoplasm of upper third of oesophagus | C153 |
| 54171 | Malignant neoplasm of middle third of oesophagus | C154 |
| 42416 | Malignant neoplasm of lower third of oesophagus | C155 |
| 67497 | Malignant neoplasm, overlapping lesion of oesophagus | C158 |
| 53591 | Malignant neoplasm of other specified part of oesophagus | C159 |
| 30700 | Malignant neoplasm of oesophagus NOS | C159 |
| 4865 | Oesophageal cancer | C159 |
| 94278 | Malignant neoplasm of gastro-oesophageal junction | C160 |
| 32022 | Malignant neoplasm of cardia of stomach | C160 |
| 22894 | Malignant neoplasm of cardio-oesophageal junction of stomach | C160 |
| 37859 | Malignant neoplasm of cardia of stomach NOS | C160 |
| 97499 | Siewert type II adenocarcinoma | C160 |
| 10368 | Gastric neoplasm | C160-C169 |
| 8386 | Malignant neoplasm of stomach | C160-C169 |
| 32362 | Malignant neoplasm of fundus of stomach | C161 |
| 43572 | Malignant neoplasm of body of stomach | C162 |
| 96094 | Siewert type III adenocarcinoma | C162 |
| 19318 | Malignant neoplasm of pyloric antrum of stomach | C163 |
| 41215 | Malignant neoplasm of pyloric canal of stomach | C164 |
| 59092 | Malignant neoplasm of pylorus of stomach NOS | C164 |
| 21620 | Malignant neoplasm of pylorus of stomach | C164 |
| 48237 | Malignant neoplasm of prepylorus of stomach | C164 |
| 42193 | Malignant neoplasm of lesser curve of stomach unspecified | C165 |
| 55434 | Malignant neoplasm of greater curve of stomach unspecified | C166 |
| 65312 | Malignant neoplasm of anterior wall of stomach NEC | C168 |
| 51690 | Malignant neoplasm, overlapping lesion of stomach | C168 |
| 96802 | Malignant neoplasm of posterior wall of stomach NEC | C168 |
| 55019 | Malignant neoplasm of other specified site of stomach | C169 |
| 65372 | Malignant neoplasm of other specified site of stomach NOS | C169 |
| 14800 | Malignant neoplasm of stomach NOS | C169 |
| 22163 | Carcinoma of caecum | C180 |
| 3811 | Malignant neoplasm of caecum | C180 |
| 1220 | Malignant neoplasm of colon | C180-C189 |
| 18632 | Malignant neoplasm of appendix | C181 |
| 10946 | Malignant neoplasm of ascending colon | C182 |
| 9088 | Malignant neoplasm of hepatic flexure of colon | C183 |
| 6935 | Malignant neoplasm of transverse colon | C184 |
| 18619 | Malignant neoplasm of splenic flexure of colon | C185 |
| 10864 | Malignant neoplasm of descending colon | C186 |
| 2815 | Malignant neoplasm of sigmoid colon | C187 |
| 93478 | Malignant neoplasm, overlapping lesion of colon | C188 |
| 9118 | Colonic cancer | C189 |
| 48231 | Malignant neoplasm of other specified sites of colon | C189 |
| 28163 | Malignant neoplasm of colon NOS | C189 |
| 27855 | Malignant neoplasm of rectosigmoid junction | C19 |
| 35357 | Malignant neoplasm of rectum, rectosigmoid junction and anus | C19-C21 |
| 7219 | Carcinoma of rectum | C20 |
| 1800 | Malignant neoplasm of rectum | C20 |
| 5901 | Rectal carcinoma | C20 |
| 50974 | Malignant neoplasm rectum, recto-sigmoid junction and anus NOS | C218 |
| 55659 | Malignant neoplasm other site rectum, rectosigmoid junction and anus | C218 |
| 45766 | [X]Malignant neoplasm of intestinal tract, part unspecified | C260 |
| 11628 | Cancer of bowel | C260 |
| 17559 | Malignant neoplasm of intestinal tract, part unspecified | C260 |
| 33444 | Malignant neoplasm of hilus of lung | C340 |
| 17391 | Malignant neoplasm of carina of bronchus | C340 |
| 12870 | Malignant neoplasm of main bronchus | C340 |
| 21698 | Malignant neoplasm of main bronchus NOS | C340 |
| 25886 | Malignant neoplasm of upper lobe of lung | C341 |
| 10358 | Malignant neoplasm of upper lobe, bronchus or lung | C341 |
| 31700 | Malignant neoplasm of upper lobe bronchus | C341 |
| 44169 | Malignant neoplasm of upper lobe, bronchus or lung NOS | C341 |
| 20170 | Pancoast's syndrome | C341 |
| 41523 | Malignant neoplasm of middle lobe bronchus | C342 |
| 54134 | Malignant neoplasm of middle lobe, bronchus or lung NOS | C342 |
| 39923 | Malignant neoplasm of middle lobe of lung | C342 |
| 31268 | Malignant neoplasm of middle lobe, bronchus or lung | C342 |
| 31188 | Malignant neoplasm of lower lobe, bronchus or lung | C343 |
| 12582 | Malignant neoplasm of lower lobe of lung | C343 |
| 42566 | Malignant neoplasm of lower lobe, bronchus or lung NOS | C343 |
| 18678 | Malignant neoplasm of lower lobe bronchus | C343 |
| 36371 | Malignant neoplasm of overlapping lesion of bronchus & lung | C348 |
| 3903 | Malignant neoplasm of bronchus or lung NOS | C349 |
| 2587 | Lung cancer | C349 |
| 40595 | [X]Malignant neoplasm of bronchus or lung, unspecified | C349 |
| 38961 | Malignant neoplasm of other sites of bronchus or lung | C349 |
| 348 | Ca female breast | C50 |
| 3968 | Malignant neoplasm of female breast | C50 |
| 26853 | Malignant neoplasm of nipple and areola of female breast | C500 |
| 64686 | Malignant neoplasm of areola of female breast | C500 |
| 59831 | Malignant neoplasm of nipple or areola of female breast NOS | C500 |
| 23380 | Malignant neoplasm of nipple of female breast | C500 |
| 12499 | [X]Malignant neoplasm of breast | C500-C509 |
| 31546 | Malignant neoplasm of central part of female breast | C501 |
| 29826 | Malignant neoplasm of upper-inner quadrant of female breast | C502 |
| 45222 | Malignant neoplasm of lower-inner quadrant of female breast | C503 |
| 23399 | Malignant neoplasm of upper-outer quadrant of female breast | C504 |
| 42070 | Malignant neoplasm of lower-outer quadrant of female breast | C505 |
| 20685 | Malignant neoplasm of axillary tail of female breast | C506 |
| 95057 | Malignant neoplasm of ectopic site of female breast | C508 |
| 49148 | Malignant neoplasm, overlapping lesion of breast | C508 |
| 9470 | Malignant neoplasm of female breast NOS | C509 |
| 56715 | Malignant neoplasm of other site of female breast | C509 |
| 38475 | Malignant neoplasm of other site of female breast NOS | C509 |
| 57235 | Malignant neoplasm of endocervical canal | C530 |
| 48820 | Malignant neoplasm of endocervix | C530 |
| 50285 | Malignant neoplasm of endocervix NOS | C530 |
| 53103 | Malignant neoplasm of endocervical gland | C530 |
| 3230 | Cervical carcinoma (uterus) | C530-C539 |
| 2747 | Malignant neoplasm of cervix uteri | C530-C539 |
| 50297 | Malignant neoplasm of exocervix | C531 |
| 58094 | Malignant neoplasm, overlapping lesion of cervix uteri | C538 |
| 95505 | Malignant neoplasm of cervical stump | C538 |
| 57719 | Malignant neoplasm of squamocolumnar junction of cervix | C538 |
| 32955 | Malignant neoplasm of other site of cervix | C539 |
| 28311 | Malignant neoplasm of cervix uteri NOS | C539 |
| 43435 | Malignant neoplasm of other site of cervix NOS | C539 |
| 1986 | Cancer of ovary | C56 |
| 7805 | Malignant neoplasm of ovary | C56 |
| 780 | Malignant neoplasm of prostate | C61 |
| 46042 | Lambda light chain myeloma | C900 |
| 43552 | Kahler's disease | C900 |
| 4944 | Multiple myeloma | C900 |
| 15211 | Myelomatosis | C900 |
| 39187 | Plasma cell leukaemia | C901 |
| 19028 | Solitary myeloma | C902 |
| 22158 | Malignant plasma cell neoplasm, extramedullary plasmacytoma | C902 |
| 21329 | Plasmacytoma NOS | C902 |

**S2: Age-specific mortality rates by sex, deprivation quintile, COVID-19 adjustment, and year (2017-2021)**

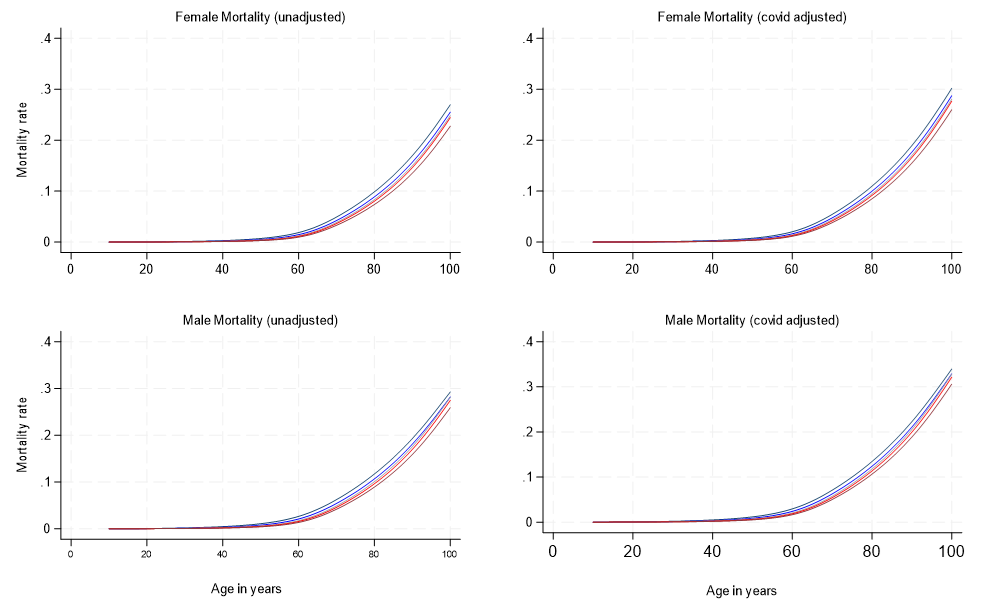

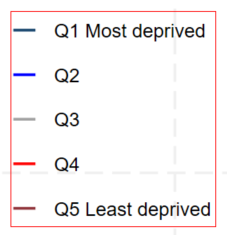

*The panel shows unadjusted (left) and COVID-adjusted (right) mortality rates for females (top) and males (bottom). Overall, mortality increased with age and was consistently higher in more deprived groups (Q1, dark blue) compared with less deprived groups (Q5, red). Adjustment for COVID-19-related deaths shifted mortality rates upward across all groups, with a greater effect in males***.**

**S3. Flow chart of participant selection form the NCRAS and the CPRD**

**Exclude (n=1,756)**

DCOs(n=471)

Missing RTD (n=3)

Aged <40years (n=345)

Male breast cancer (n=263)

Female prostate (n=1)

IMD missing (78)

Ethnicity missing (n=589)

Invalid dates (n=6)

Total eligible

N=247,512

Eligible from NCRAS

n=160,956

**CPRD (2017-2019)**

**N=86,877**

**NCRAS (2006-2016)**

**N=** **162,712**

**Exclude (n=321)**

Male breast cancer (n=174)

Female prostate (n=4)

Invalid dates (n=61)

IMD missing (n=82)

Eligible from CPRD

n= 86,556

**Excluded**

Aged>100years at

diagnosis (n=84)

**Included for analysis**

**(n=247,428)**

*DCO=Death Certificate Only; RTD=Route to Diagnosis; IMD=Index of multiple deprivation; invalid dates=where recorded date of death precedes the date of cancer diagnosis. Our final selection included* ***160,885*** *patients from the NCRAS cohort - diagnosed between 2006 and 2016 – and* ***86,543*** *extracted from the CPRD - diagnosed between 2017 and 2019.*

**S4. Proportion who died during follow-up by ethnicity and cancer type**

Proportion of deaths during follow-up

|  | All ethnicities | White | Black | Asian | Mixed | Other |
| --- | --- | --- | --- | --- | --- | --- |
| Breast | 17.95 | 18.59 | 16.42 | 10.01 | 12.68 | 13.69 |
| Lung | 82.73 | 83.13 | 74.91 | 68.77 | 75.25 | 89.38 |
| Prostate | 22.38 | 23.28 | 13.4 | 13.94 | 16.12 | 18.32 |
| CRC | 47.98 | 48.51 | 44.85 | 36.71 | 38.19 | 50.15 |
| OG | 81.86 | 82.43 | 70.67 | 64.09 | 75.37 | 88.35 |
| Cervix | 34.01 | 34.36 | 36.54 | 30.14 | 23.88 | 37.04 |
| Myeloma | 49.5 | 51.63 | 33.77 | 39.13 | 36.02 | 38.46 |
| Ovary | 53.87 | 54.9 | 56.86 | 37.19 | 34.08 | 57.79 |
| All site | 43.11 | 44.22 | 28.77 | 27.96 | 32.9 | 42.76 |

*Figure show the proportion of deaths recorded during follow‑up (2010-2021) across major cancer sites, stratified by ethnic group. Categories (from left to right) include: All ethnicities, White, Black, Asian, Mixed, and Other ethnic group. Error bars = 95% confidence intervals; OG =Oesophagogastric; error bars=95% confidence interval*

**S5. Ethnic differences in net survival by age and cancer type**

| **Breast cancer** | | | | | | | | | | | | | | | | | | | | | |
| --- | --- | --- | --- | --- | --- | --- | --- | --- | --- | --- | --- | --- | --- | --- | --- | --- | --- | --- | --- | --- | --- |
|  |  | **All ages** | | | | **40-59** | | | | **60-69** | | | | **70-79** | | | | **Over 80s** | | | |
| **Ethnicity** | **Follow-up time** | **Beginning total** | **Net Survival** | **Lower limit** | **Upper limit** | **Beginning total** | **Net Survival** | **Lower limit** | **Upper limit** | **Beginning total** | **Net Survival** | **Lower limit** | **Upper limit** | **Beginning total** | **Net Survival** | **Lower limit** | **Upper limit** | **Beginning total** | **Net Survival** | **Lower limit** | **Upper limit** |
| All | 1 | 62962 | 95.34 | 95.18 | 95.51 | 26164 | 98.72 | 98.58 | 98.86 | 16044 | 97.79 | 97.56 | 98.02 | 11227 | 94.51 | 94.09 | 94.93 | 9527 | 82.92 | 82.17 | 83.68 |
|  | 3 | 60026 | 87.21 | 86.94 | 87.48 | 25829 | 94.77 | 94.49 | 95.05 | 15689 | 93.11 | 92.7 | 93.51 | 10610 | 84.69 | 83.99 | 85.38 | 7898 | 59.42 | 58.39 | 60.45 |
|  | 5 | 40262 | 80.41 | 80.04 | 80.78 | 18170 | 91.09 | 90.67 | 91.51 | 11165 | 88.89 | 88.31 | 89.47 | 6776 | 75.84 | 74.87 | 76.82 | 4151 | 42.07 | 40.83 | 43.31 |
| White | 1 | 55986 | 95.19 | 95.01 | 95.36 | 22328 | 98.7 | 98.55 | 98.85 | 14436 | 97.81 | 97.57 | 98.05 | 10263 | 94.45 | 94.01 | 94.89 | 8959 | 83.04 | 82.27 | 83.82 |
|  | 3 | 53288 | 86.88 | 86.59 | 87.17 | 22038 | 94.86 | 94.56 | 95.16 | 14119 | 93.11 | 92.68 | 93.54 | 9693 | 84.63 | 83.91 | 85.36 | 7438 | 59.41 | 58.35 | 60.47 |
|  | 5 | 36163 | 79.95 | 79.56 | 80.34 | 15796 | 91.15 | 90.7 | 91.6 | 10186 | 88.8 | 88.19 | 89.41 | 6254 | 75.87 | 74.86 | 76.88 | 3927 | 42.07 | 40.8 | 43.34 |
| Black | 1 | 1394 | 95.91 | 94.87 | 96.95 | 912 | 97.81 | 96.86 | 98.76 | 223 | 95.97 | 93.39 | 98.54 | 170 | 92.36 | 88.38 | 96.34 | 89 | 83.17 | 75.45 | 90.88 |
|  | 3 | 1337 | 86.53 | 84.65 | 88.41 | 892 | 89.81 | 87.76 | 91.87 | 214 | 87.97 | 83.4 | 92.53 | 157 | 80.84 | 74.75 | 86.94 | 74 | 59.6 | 48.8 | 70.4 |
|  | 5 | 805 | 81.04 | 78.55 | 83.53 | 546 | 85.6 | 82.83 | 88.37 | 125 | 85.21 | 79.82 | 90.59 | 103 | 69.75 | 61.8 | 77.7 | 31 | 43.97 | 28.12 | 59.82 |
| Asian | 1 | 2125 | 97.84 | 97.22 | 98.46 | 1240 | 99.27 | 98.8 | 99.75 | 517 | 98.26 | 97.13 | 99.39 | 240 | 96.26 | 93.86 | 98.65 | 128 | 85.17 | 79.05 | 91.3 |
|  | 3 | 2079 | 92.88 | 91.72 | 94.04 | 1231 | 96.2 | 95.06 | 97.34 | 508 | 93.95 | 91.8 | 96.1 | 231 | 87.68 | 83.28 | 92.09 | 109 | 66.07 | 57.33 | 74.81 |
|  | 5 | 1305 | 86.96 | 85.07 | 88.85 | 800 | 91.28 | 89.13 | 93.43 | 321 | 89.74 | 86.28 | 93.2 | 134 | 76.35 | 68.89 | 83.81 | 50 | 50.63 | 39.3 | 61.96 |
| Mixed | 1 | 2128 | 97.09 | 96.38 | 97.8 | 1036 | 99.04 | 98.44 | 99.63 | 506 | 99.21 | 98.44 | 99.98 | 369 | 96.75 | 94.95 | 98.56 | 217 | 83.43 | 78.49 | 88.37 |
|  | 3 | 2066 | 90.33 | 88.98 | 91.67 | 1026 | 94.84 | 93.36 | 96.31 | 502 | 96.17 | 94.42 | 97.91 | 357 | 85.48 | 81.61 | 89.34 | 181 | 63.91 | 57.17 | 70.65 |
|  | 5 | 1172 | 83.19 | 80.89 | 85.49 | 598 | 92.27 | 90.08 | 94.46 | 297 | 91.7 | 88.05 | 95.35 | 185 | 74.29 | 67.64 | 80.94 | 92 | 39.95 | 30.75 | 49.14 |
| Other | 1 | 1329 | 94.51 | 93.29 | 95.73 | 648 | 99.07 | 98.34 | 99.81 | 362 | 95.58 | 93.47 | 97.7 | 185 | 92.98 | 89.3 | 96.65 | 134 | 71.66 | 64.06 | 79.26 |
|  | 3 | 1256 | 88.05 | 86.23 | 89.86 | 642 | 95.69 | 94.03 | 97.36 | 346 | 90.75 | 87.68 | 93.81 | 172 | 86.05 | 80.83 | 91.28 | 96 | 47.6 | 38.92 | 56.29 |
|  | 5 | 817 | 85.61 | 83.48 | 87.74 | 430 | 94.85 | 92.81 | 96.89 | 236 | 89.92 | 86.67 | 93.17 | 100 | 81.06 | 74.54 | 87.58 | 51 | 37.81 | 28.43 | 47.19 |
|  | | **Lung cancer** | | | | | | | | | | | | | | | | | | | |
| All | 1 | 45698 | 39.48 | 39.04 | 39.93 | 5935 | 48.66 | 47.39 | 49.93 | 12419 | 45.34 | 44.46 | 46.21 | 15703 | 39.93 | 39.16 | 40.69 | 11641 | 27.97 | 27.15 | 28.78 |
|  | 3 | 18042 | 20.02 | 19.64 | 20.39 | 2888 | 27.76 | 26.58 | 28.93 | 5630 | 24.97 | 24.19 | 25.75 | 6269 | 19.96 | 19.31 | 20.61 | 3255 | 10.82 | 10.23 | 11.41 |
|  | 5 | 6413 | 14.4 | 14.01 | 14.78 | 1159 | 21.74 | 20.53 | 22.95 | 2251 | 19.48 | 18.67 | 20.29 | 2131 | 13.71 | 13.03 | 14.38 | 872 | 5.78 | 5.21 | 6.35 |
| White | 1 | 42393 | 39.28 | 38.81 | 39.74 | 5237 | 48.18 | 46.83 | 49.53 | 11497 | 45.1 | 44.19 | 46.01 | 14651 | 40.14 | 39.34 | 40.93 | 11008 | 27.82 | 26.99 | 28.66 |
|  | 3 | 16650 | 19.69 | 19.3 | 20.08 | 2523 | 27.06 | 25.82 | 28.29 | 5185 | 24.65 | 23.85 | 25.46 | 5880 | 19.91 | 19.24 | 20.59 | 3062 | 10.65 | 10.05 | 11.26 |
|  | 5 | 5915 | 14.13 | 13.73 | 14.52 | 1018 | 21.24 | 19.98 | 22.51 | 2085 | 19.19 | 18.36 | 20.02 | 1995 | 13.64 | 12.95 | 14.34 | 817 | 5.68 | 5.1 | 6.26 |
| Black | 1 | 535 | 46.73 | 42.51 | 50.95 | 215 | 47.91 | 41.26 | 54.56 | 114 | 51.76 | 42.65 | 60.86 | 133 | 47.37 | 38.96 | 55.79 | 73 | 34.26 | 23.52 | 44.99 |
|  | 3 | 250 | 26.7 | 22.77 | 30.64 | 103 | 27.26 | 20.99 | 33.54 | 59 | 31.41 | 22.48 | 40.33 | 63 | 26.28 | 18.45 | 34.12 | 25 | 17.82 | 9.24 | 26.4 |
|  | 5 | 89 | 20.55 | 16.05 | 25.04 | 32 | 22.81 | 15.81 | 29.82 | 23 | 29.31 | 20.14 | 38.49 | 23 | 17.56 | 8.93 | 26.18 | 11 | 9 | 0.96 | 17.05 |
| Asian | 1 | 716 | 53.77 | 50.13 | 57.42 | 174 | 61.49 | 54.3 | 68.69 | 195 | 61.54 | 54.74 | 68.34 | 216 | 50.93 | 44.3 | 57.56 | 131 | 36.65 | 28.47 | 44.83 |
|  | 3 | 385 | 33.71 | 30.12 | 37.3 | 107 | 39.81 | 32.35 | 47.28 | 120 | 40.08 | 32.9 | 47.26 | 110 | 30.13 | 23.82 | 36.44 | 48 | 22.2 | 15 | 29.4 |
|  | 5 | 162 | 26.42 | 22.49 | 30.34 | 49 | 31.61 | 23.77 | 39.45 | 49 | 33.1 | 24.96 | 41.25 | 45 | 22.66 | 15.72 | 29.6 | 19 | 15.7 | 7.87 | 23.53 |
| Mixed | 1 | 1178 | 46.27 | 43.43 | 49.11 | 170 | 54.71 | 47.26 | 62.15 | 339 | 56.64 | 51.38 | 61.9 | 418 | 42.11 | 37.39 | 46.83 | 251 | 33.48 | 27.67 | 39.28 |
|  | 3 | 545 | 26.82 | 24.14 | 29.49 | 93 | 37.79 | 30.24 | 45.33 | 192 | 36.18 | 30.8 | 41.57 | 176 | 23.24 | 18.99 | 27.5 | 84 | 12.62 | 8.16 | 17.07 |
|  | 5 | 189 | 18.76 | 15.64 | 21.89 | 41 | 24.3 | 15.29 | 33.3 | 72 | 28.81 | 22.63 | 34.98 | 57 | 15.86 | 10.81 | 20.92 | 19 | 5.99 | 1.4 | 10.58 |
| Other | 1 | 876 | 24.2 | 21.38 | 27.03 | 139 | 44.6 | 36.4 | 52.81 | 274 | 27.01 | 21.79 | 32.23 | 285 | 14.04 | 10.04 | 18.03 | 178 | 20.23 | 14.39 | 26.07 |
|  | 3 | 212 | 11.44 | 9.25 | 13.63 | 62 | 27.69 | 20.06 | 35.33 | 74 | 11.18 | 7.36 | 15.01 | 40 | 6.79 | 3.78 | 9.81 | 36 | 6.64 | 2.81 | 10.46 |
|  | 5 | 58 | 8.26 | 5.9 | 10.61 | 19 | 21.1 | 11.62 | 30.58 | 22 | 8.37 | 4.63 | 12.11 | 11 | 4.95 | 1.97 | 7.94 | 6 | 3.69 | 0.26 | 7.12 |
| **Prostate cancer** | | | | | | | | | | | | | | | | | | | | | |
| All | 1 | 58847 | 94.32 | 94.13 | 94.5 | 7579 | 98.84 | 98.6 | 99.08 | 19522 | 98.09 | 97.9 | 98.28 | 21717 | 95.57 | 95.3 | 95.84 | 10029 | 80.84 | 80.07 | 81.61 |
|  | 3 | 55498 | 83.99 | 83.68 | 84.3 | 7491 | 95.25 | 94.74 | 95.75 | 19149 | 92.81 | 92.43 | 93.18 | 20753 | 85.56 | 85.07 | 86.05 | 8105 | 54.9 | 53.88 | 55.91 |
|  | 5 | 35015 | 75.63 | 75.2 | 76.05 | 4977 | 92.33 | 91.59 | 93.07 | 13140 | 88.09 | 87.53 | 88.64 | 12969 | 76.3 | 75.58 | 77.01 | 3929 | 37.16 | 35.94 | 38.38 |
| White | 1 | 52332 | 94.08 | 93.88 | 94.29 | 5997 | 98.78 | 98.51 | 99.06 | 17308 | 98.09 | 97.89 | 98.29 | 19693 | 95.55 | 95.26 | 95.84 | 9334 | 80.55 | 79.74 | 81.35 |
|  | 3 | 49232 | 83.42 | 83.09 | 83.75 | 5924 | 95.07 | 94.5 | 95.64 | 16977 | 92.77 | 92.37 | 93.18 | 18815 | 85.35 | 84.83 | 85.87 | 7516 | 54.41 | 53.36 | 55.47 |
|  | 5 | 31330 | 74.86 | 74.41 | 75.31 | 4033 | 92.16 | 91.33 | 92.98 | 11862 | 88 | 87.42 | 88.59 | 11801 | 75.89 | 75.14 | 76.65 | 3634 | 36.52 | 35.26 | 37.77 |
| Black | 1 | 2357 | 97.16 | 96.49 | 97.83 | 844 | 99.53 | 99.06 | 99.99 | 707 | 98.31 | 97.35 | 99.26 | 587 | 96.94 | 95.55 | 98.34 | 219 | 84.95 | 80.23 | 89.68 |
|  | 3 | 2290 | 90.65 | 89.41 | 91.88 | 840 | 96.01 | 94.57 | 97.45 | 695 | 94.87 | 93.18 | 96.56 | 569 | 88.66 | 86 | 91.32 | 186 | 62.27 | 55.57 | 68.97 |
|  | 5 | 1435 | 85.05 | 83.2 | 86.89 | 510 | 92.84 | 90.48 | 95.2 | 441 | 91.67 | 88.97 | 94.37 | 390 | 81.82 | 78.19 | 85.45 | 94 | 47.05 | 38.49 | 55.6 |
| Asian | 1 | 1086 | 96.97 | 95.95 | 97.99 | 174 | 98.85 | 97.27 | 100.43 | 407 | 99.27 | 98.44 | 100.1 | 373 | 95.99 | 94 | 97.98 | 132 | 90.17 | 85.11 | 95.24 |
|  | 3 | 1053 | 89.94 | 88.05 | 91.84 | 172 | 95.71 | 92.6 | 98.83 | 404 | 94.61 | 92.3 | 96.92 | 358 | 89.04 | 85.71 | 92.36 | 119 | 70.62 | 62.3 | 78.93 |
|  | 5 | 657 | 85.21 | 82.61 | 87.8 | 110 | 95.72 | 92.61 | 98.83 | 256 | 92.58 | 89.54 | 95.62 | 227 | 83.6 | 79.11 | 88.08 | 64 | 53.63 | 42.18 | 65.07 |
| Mixed | 1 | 1859 | 95.92 | 95.02 | 96.82 | 302 | 99.34 | 98.43 | 100.25 | 669 | 97.91 | 96.83 | 98.99 | 683 | 95.76 | 94.25 | 97.27 | 205 | 84.9 | 80.01 | 89.79 |
|  | 3 | 1783 | 87.72 | 86.12 | 89.32 | 300 | 96.48 | 94.2 | 98.76 | 655 | 91.93 | 89.68 | 94.19 | 654 | 88.34 | 85.77 | 90.92 | 174 | 59.67 | 52.57 | 66.77 |
|  | 5 | 960 | 78.59 | 75.77 | 81.42 | 169 | 91.78 | 86.51 | 97.06 | 360 | 84.61 | 80.6 | 88.63 | 347 | 76.14 | 70.86 | 81.43 | 84 | 49.6 | 41.24 | 57.97 |
| Other | 1 | 1213 | 93.99 | 92.65 | 95.33 | 262 | 97.33 | 95.38 | 99.28 | 431 | 96.99 | 95.37 | 98.6 | 381 | 93.71 | 91.27 | 96.14 | 139 | 79.16 | 72.43 | 85.88 |
|  | 3 | 1140 | 85.08 | 82.98 | 87.18 | 255 | 95.12 | 92.43 | 97.82 | 418 | 90.39 | 87.44 | 93.33 | 357 | 83.83 | 79.93 | 87.73 | 110 | 53.94 | 45.47 | 62.41 |
|  | 5 | 633 | 79.27 | 76.3 | 82.24 | 155 | 91.81 | 86.99 | 96.63 | 221 | 86.55 | 82.31 | 90.8 | 204 | 80.29 | 75.58 | 84.99 | 53 | 35.63 | 25.23 | 46.03 |
| **Colorectal cancer** | | | | | | | | | | | | | | | | | | | | | |
| All | 1 | 45319 | 75.84 | 75.45 | 76.24 | 8154 | 86.42 | 85.68 | 87.17 | 11474 | 84.72 | 84.07 | 85.38 | 13513 | 77.62 | 76.92 | 78.32 | 12178 | 58.42 | 57.54 | 59.29 |
|  | 3 | 34368 | 58.27 | 57.81 | 58.74 | 7047 | 70.49 | 69.47 | 71.52 | 9721 | 70.42 | 69.57 | 71.28 | 10488 | 60.81 | 59.96 | 61.66 | 7112 | 35.85 | 34.97 | 36.73 |
|  | 5 | 19673 | 49.8 | 49.28 | 50.33 | 4192 | 62.79 | 61.57 | 64 | 6136 | 63.53 | 62.54 | 64.51 | 6120 | 51.99 | 51.02 | 52.95 | 3225 | 25.68 | 24.76 | 26.61 |
| White | 1 | 41503 | 75.71 | 75.3 | 76.12 | 7061 | 86.46 | 85.66 | 87.26 | 10432 | 84.91 | 84.23 | 85.6 | 12511 | 77.8 | 77.07 | 78.53 | 11499 | 58.49 | 57.59 | 59.39 |
|  | 3 | 31420 | 58.02 | 57.53 | 58.51 | 6105 | 70.32 | 69.22 | 71.41 | 8858 | 70.44 | 69.55 | 71.34 | 9733 | 60.94 | 60.06 | 61.82 | 6724 | 36.01 | 35.1 | 36.91 |
|  | 5 | 18073 | 49.41 | 48.87 | 49.96 | 3666 | 62.39 | 61.09 | 63.69 | 5639 | 63.49 | 62.46 | 64.52 | 5697 | 51.98 | 50.98 | 52.99 | 3071 | 25.67 | 24.73 | 26.62 |
| Black | 1 | 682 | 79.48 | 76.45 | 82.51 | 252 | 81.75 | 76.99 | 86.5 | 139 | 87.77 | 82.35 | 93.2 | 166 | 80.13 | 74.08 | 86.17 | 125 | 64.82 | 56.49 | 73.14 |
|  | 3 | 542 | 58.02 | 54.14 | 61.9 | 206 | 66.16 | 60.04 | 72.28 | 122 | 66.71 | 58.45 | 74.97 | 133 | 53.96 | 46.12 | 61.8 | 81 | 37.57 | 28.75 | 46.39 |
|  | 5 | 279 | 51.13 | 46.71 | 55.54 | 118 | 60.31 | 53.03 | 67.59 | 63 | 55.5 | 44.84 | 66.17 | 67 | 49.14 | 40.94 | 57.35 | 31 | 29.95 | 20.7 | 39.21 |
| Asian | 1 | 933 | 82.43 | 79.99 | 84.87 | 319 | 88.72 | 85.25 | 92.18 | 277 | 88.09 | 84.28 | 91.9 | 209 | 78 | 72.4 | 83.6 | 128 | 61.73 | 53.36 | 70.1 |
|  | 3 | 769 | 68.01 | 64.92 | 71.1 | 283 | 73.9 | 68.91 | 78.89 | 244 | 77.04 | 71.89 | 82.2 | 163 | 65.86 | 59.34 | 72.37 | 79 | 37.3 | 28.54 | 46.05 |
|  | 5 | 452 | 60.45 | 56.77 | 64.14 | 167 | 69.58 | 63.91 | 75.24 | 144 | 69.29 | 62.34 | 76.24 | 108 | 52.07 | 44.28 | 59.86 | 33 | 34.54 | 25.63 | 43.45 |
| Mixed | 1 | 1212 | 81.36 | 79.17 | 83.55 | 278 | 91.37 | 88.07 | 94.66 | 346 | 84.11 | 80.26 | 87.95 | 348 | 85.06 | 81.32 | 88.8 | 240 | 60.43 | 54.27 | 66.6 |
|  | 3 | 986 | 64.95 | 62.15 | 67.75 | 254 | 77.06 | 71.85 | 82.26 | 291 | 72.04 | 67.14 | 76.94 | 296 | 69.45 | 64.43 | 74.48 | 145 | 35.12 | 28.92 | 41.31 |
|  | 5 | 530 | 57.19 | 53.77 | 60.61 | 127 | 66.81 | 59.13 | 74.49 | 168 | 64.74 | 58.28 | 71.21 | 173 | 63.59 | 57.87 | 69.32 | 62 | 26.83 | 20.24 | 33.43 |
| Other | 1 | 989 | 65.83 | 62.88 | 68.78 | 244 | 81.56 | 76.7 | 86.41 | 280 | 73.57 | 68.42 | 78.72 | 279 | 58.43 | 52.66 | 64.19 | 186 | 44.64 | 37.53 | 51.74 |
|  | 3 | 651 | 52.04 | 48.83 | 55.24 | 199 | 68.48 | 62.48 | 74.47 | 206 | 63.08 | 57.28 | 68.88 | 163 | 44.41 | 38.39 | 50.42 | 83 | 24.94 | 18.42 | 31.45 |
|  | 5 | 339 | 47.97 | 44.47 | 51.48 | 114 | 64.21 | 57.25 | 71.18 | 122 | 61.15 | 55.12 | 67.18 | 75 | 41.46 | 35.18 | 47.74 | 28 | 15.41 | 8.49 | 22.33 |
| **Oesophagogastric cancer** | | | | | | | | | | | | | | | | | | | | | |
| All | 1 | 16281 | 44.53 | 43.77 | 45.29 | 2682 | 58.05 | 56.19 | 59.92 | 4082 | 53.11 | 51.58 | 54.64 | 4961 | 44.73 | 43.35 | 46.11 | 4556 | 28.65 | 27.34 | 29.96 |
|  | 3 | 7249 | 20.91 | 20.26 | 21.55 | 1557 | 31.11 | 29.32 | 32.91 | 2168 | 26.4 | 25.01 | 27.79 | 2219 | 21.44 | 20.27 | 22.62 | 1305 | 9.36 | 8.48 | 10.24 |
|  | 5 | 2555 | 16.02 | 15.38 | 16.66 | 630 | 26.84 | 25 | 28.68 | 810 | 21.24 | 19.84 | 22.64 | 802 | 15.78 | 14.62 | 16.94 | 313 | 5.01 | 4.19 | 5.82 |
| White | 1 | 14956 | 44.29 | 43.5 | 45.09 | 2364 | 57.57 | 55.58 | 59.56 | 3741 | 53.12 | 51.52 | 54.71 | 4575 | 44.97 | 43.53 | 46.41 | 4276 | 28.52 | 27.16 | 27.16 |
|  | 3 | 6624 | 20.48 | 19.82 | 21.15 | 1361 | 29.79 | 27.9 | 31.68 | 1987 | 26.03 | 24.58 | 27.47 | 2057 | 21.61 | 20.38 | 22.83 | 1219 | 9.27 | 8.37 | 8.37 |
|  | 5 | 2316 | 15.51 | 14.85 | 16.16 | 534 | 25.49 | 23.56 | 27.42 | 742 | 20.64 | 19.19 | 22.08 | 746 | 15.83 | 14.62 | 17.03 | 294 | 5.01 | 4.19 | 4.19 |
| Black | 1 | 277 | 54.88 | 49.04 | 60.72 | 90 | 67.78 | 58.19 | 77.36 | 47 | 55.32 | 41.32 | 69.32 | 71 | 46.48 | 35.01 | 57.95 | 69 | 46.39 | 34.76 | 34.76 |
|  | 3 | 152 | 29.12 | 23.53 | 34.71 | 61 | 47.67 | 37.19 | 58.14 | 26 | 37.65 | 23.33 | 51.96 | 33 | 12.88 | 5.1 | 20.67 | 32 | 15.63 | 6.31 | 6.31 |
|  | 5 | 58 | 26.01 | 20.25 | 31.76 | 32 | 47.67 | 37.19 | 58.14 | 12 | 29.34 | 14.53 | 44.14 | 8 | 10.31 | 2.88 | 17.73 | 6 | 0 | 0 | 0 |
| Asian | 1 | 295 | 57.29 | 51.66 | 62.92 | 88 | 67.05 | 57.3 | 76.79 | 81 | 62.97 | 52.53 | 73.4 | 75 | 53.34 | 42.17 | 64.51 | 51 | 37.26 | 24.25 | 24.25 |
|  | 3 | 169 | 38.56 | 32.94 | 44.18 | 59 | 48.84 | 38.49 | 59.19 | 51 | 41.1 | 29.99 | 52.21 | 40 | 32.33 | 21.7 | 42.95 | 19 | 25.02 | 13.37 | 13.37 |
|  | 5 | 82 | 33.01 | 26.94 | 39.08 | 38 | 39.64 | 28.43 | 50.85 | 18 | 41.1 | 29.99 | 52.22 | 18 | 25.12 | 14.22 | 36.02 | 8 | 25.02 | 13.37 | 13.37 |
| Mixed | 1 | 398 | 52.26 | 47.37 | 57.16 | 73 | 65.75 | 54.97 | 76.54 | 108 | 64.82 | 55.87 | 73.77 | 139 | 47.49 | 39.24 | 55.73 | 78 | 30.78 | 20.68 | 20.68 |
|  | 3 | 208 | 25.49 | 21.06 | 29.93 | 48 | 43.92 | 32.07 | 55.78 | 70 | 33.03 | 23.85 | 42.21 | 66 | 21.97 | 15.04 | 28.9 | 24 | 4.89 | 0.19 | 0.19 |
|  | 5 | 68 | 20 | 15.21 | 24.8 | 20 | 38.39 | 25.9 | 50.87 | 22 | 30.97 | 21.56 | 40.37 | 23 | 15.41 | 8.02 | 22.8 | 3 | 0 | 0 | 0 |
| Other | 1 | 355 | 27.04 | 22.45 | 31.64 | 67 | 41.79 | 30.16 | 53.43 | 105 | 32.38 | 23.53 | 41.23 | 101 | 22.77 | 14.71 | 30.84 | 82 | 13.42 | 6.23 | 6.23 |
|  | 3 | 96 | 12.51 | 9.02 | 15.99 | 28 | 17.51 | 7.96 | 27.07 | 34 | 16.9 | 9.81 | 24 | 23 | 11.67 | 5.52 | 17.82 | 11 | 3.66 | 0 | 0.08 |
|  | 5 | 31 | 11.64 | 8.21 | 15.08 | 6 | 17.51 | 7.96 | 27.07 | 16 | 16.91 | 9.81 | 24 | 7 | 11.67 | 5.53 | 17.82 | 2 | 0 | 0 | 0 |
| **Myeloma** | | | | | | | | | | | | | | | | | | | | | |
| All | 1 | 6559 | 79.56 | 78.59 | 80.54 | 1231 | 92.69 | 91.24 | 94.14 | 1585 | 86.75 | 85.09 | 88.42 | 2044 | 81.61 | 79.93 | 83.29 | 1699 | 60.87 | 58.56 | 63.19 |
|  | 3 | 5218 | 60.08 | 58.84 | 61.31 | 1141 | 82.7 | 80.5 | 84.9 | 1375 | 71.66 | 69.35 | 73.97 | 1668 | 59.23 | 57 | 61.46 | 1034 | 33.61 | 31.23 | 35.99 |
|  | 5 | 2768 | 44.37 | 42.82 | 45.91 | 715 | 73.04 | 69.89 | 76.2 | 816 | 55.41 | 52.27 | 58.55 | 851 | 40.68 | 37.87 | 43.48 | 386 | 16.81 | 14.24 | 19.37 |
| White | 1 | 5656 | 78.53 | 77.46 | 79.59 | 944 | 92.06 | 90.33 | 93.78 | 1375 | 86.33 | 84.51 | 88.14 | 1799 | 81.27 | 79.47 | 83.08 | 1538 | 60.03 | 57.58 | 62.47 |
|  | 3 | 4441 | 58.52 | 57.19 | 59.86 | 869 | 81.95 | 79.4 | 84.49 | 1187 | 71.05 | 68.56 | 73.54 | 1462 | 58.36 | 55.97 | 60.75 | 923 | 32.86 | 30.39 | 35.34 |
|  | 5 | 2362 | 42.68 | 41.04 | 44.33 | 553 | 72.09 | 68.54 | 75.64 | 722 | 55.07 | 51.76 | 58.39 | 739 | 39.45 | 36.47 | 42.42 | 348 | 16.36 | 13.76 | 18.95 |
| Black | 1 | 384 | 89.07 | 85.95 | 92.19 | 152 | 95.4 | 92.07 | 98.72 | 72 | 88.89 | 81.7 | 96.09 | 96 | 87.51 | 80.93 | 94.09 | 64 | 76.58 | 66.29 | 86.87 |
|  | 3 | 342 | 73.01 | 68.36 | 77.66 | 145 | 84.87 | 78.88 | 90.86 | 64 | 79.23 | 69.51 | 88.95 | 84 | 65.59 | 55.72 | 75.47 | 49 | 49.12 | 36.4 | 61.83 |
|  | 5 | 192 | 59.84 | 53.41 | 66.28 | 93 | 76.89 | 67.7 | 86.09 | 34 | 56.72 | 39.82 | 73.62 | 47 | 47.77 | 35.91 | 59.64 | 18 | 38.67 | 22.01 | 55.33 |
| Asian | 1 | 206 | 87.87 | 83.42 | 92.32 | 71 | 91.55 | 85.13 | 97.97 | 57 | 96.49 | 91.76 | 101.23 | 46 | 84.79 | 74.56 | 95.02 | 32 | 68.76 | 53 | 53 |
|  | 3 | 181 | 72.23 | 65.89 | 78.57 | 65 | 82.24 | 73.16 | 91.33 | 55 | 80.6 | 69.85 | 91.35 | 39 | 64.62 | 50.83 | 78.4 | 22 | 45.54 | 27.55 | 27.55 |
|  | 5 | 109 | 55.31 | 46.72 | 63.89 | 44 | 71.97 | 58.24 | 85.69 | 30 | 63.57 | 47.98 | 79.17 | 24 | 46.62 | 30.02 | 63.22 | 11 | 0 | 0 | 0 |
| Mixed | 1 | 210 | 84.29 | 79.38 | 89.2 | 46 | 97.83 | 93.66 | 101.99 | 55 | 89.09 | 80.93 | 97.25 | 67 | 83.59 | 74.79 | 92.39 | 42 | 64.3 | 50.02 | 50.02 |
|  | 3 | 177 | 64.89 | 57.82 | 71.96 | 45 | 87.94 | 78.06 | 97.83 | 49 | 70.6 | 58.08 | 83.12 | 56 | 65.37 | 53.21 | 77.53 | 27 | 31.74 | 15.17 | 15.17 |
|  | 5 | 69 | 49.44 | 37.65 | 61.22 | 18 | 80.61 | 64.63 | 96.6 | 19 | 45.39 | 18.54 | 72.24 | 26 | 50.63 | 33.06 | 68.2 | 6 | 0 | 0 | 0 |
| Other | 1 | 103 | 74.76 | 66.43 | 83.09 | 18 | 94.44 | 84.16 | 104.73 | 26 | 76.93 | 61.23 | 92.62 | 36 | 75 | 61.09 | 88.92 | 23 | 56.53 | 37.04 | 37.04 |
|  | 3 | 77 | 62.72 | 52.78 | 72.65 | 17 | 94.45 | 84.16 | 104.73 | 20 | 69.24 | 52.01 | 86.46 | 27 | 67.66 | 51.88 | 83.44 | 13 | 23.2 | 4.62 | 4.62 |
|  | 5 | 36 | 54.06 | 40.94 | 67.18 | 7 | 78.71 | 51.61 | 105.81 | 11 | 62.95 | 43.69 | 82.21 | 15 | 67.67 | 51.89 | 83.45 | 3 | 0 | 0 | 0 |

| **Ovarian cancer** | | | | | | | | | | | | | | | | | | | | | | | | | | | | | | | |
| --- | --- | --- | --- | --- | --- | --- | --- | --- | --- | --- | --- | --- | --- | --- | --- | --- | --- | --- | --- | --- | --- | --- | --- | --- | --- | --- | --- | --- | --- | --- | --- |
|  | |  | **All ages** | | | | | | | | **40-59** | | | | | | | | **60-69** | | | | | | | | **Over 70s** | | | | |
| **Ethnicity** | | **Follow-up time** | **Beginning total** | | **Net Survival** | | **Lower limit** | | **Upper limit** | | **Beginning total** | | **Net Survival** | | **Lower limit** | | **Upper limit** | | **Beginning total** | | **Net Survival** | | **Lower limit** | | **Upper limit** | **Beginning total** | | | **Net Survival** | **Lower limit** | **Upper limit** |
| All | | 1 | 7036 | | 73.55 | | 72.52 | | 74.58 | | 2297 | | 91.12 | | 89.96 | | 92.28 | | 1794 | | 81.38 | | 79.58 | | 83.18 | 2945 | | | 55.08 | 53.29 | 56.88 |
|  |  | 3 | 5175 | | 52.81 | | 51.62 | | 54.01 | | 2093 | | 75.85 | | 74.03 | | 77.66 | | 1460 | | 55.39 | | 53.02 | | 57.76 | 1622 | | | 33.31 | 31.56 | 35.06 |
|  |  | 5 | 2789 | | 43.38 | | 42.06 | | 44.7 | | 1306 | | 66.3 | | 64.05 | | 68.55 | | 774 | | 44.38 | | 41.76 | | 47 | 709 | | | 24.81 | 22.98 | 26.63 |
| White | | 1 | 6367 | | 72.93 | | 71.84 | | 74.02 | | 1996 | | 90.93 | | 89.67 | | 92.19 | | 1614 | | 81.72 | | 79.84 | | 79.84 | 1618 | | | 66.51 | 64.21 | 68.8 |
|  |  | 3 | 4643 | | 51.99 | | 50.73 | | 53.25 | | 1815 | | 75.7 | | 73.76 | | 77.64 | | 1319 | | 55.71 | | 53.22 | | 53.22 | 1076 | | | 42.6 | 40.11 | 45.09 |
|  |  | 5 | 2523 | | 42.71 | | 41.33 | | 44.08 | | 1152 | | 66.57 | | 64.19 | | 68.96 | | 714 | | 44.51 | | 41.77 | | 41.77 | 505 | | | 32.36 | 29.66 | 35.05 |
| Black | | 1 | 101 | | 74.26 | | 65.79 | | 82.73 | | 48 | | 87.5 | | 78.25 | | 96.75 | | 19 | | 68.42 | | 48.19 | | 48.19 | 25 | | | 64 | 45.66 | 82.35 |
|  |  | 3 | 75 | | 48.82 | | 38.66 | | 58.97 | | 42 | | 59.23 | | 45.18 | | 73.27 | | 13 | | 41.06 | | 18.13 | | 18.13 | 16 | | | 49.93 | 30.13 | 69.73 |
|  |  | 5 | 31 | | 38.76 | | 28.02 | | 49.49 | | 21 | | 41.74 | | 26.59 | | 56.88 | | 2 | | 41.06 | | 18.14 | | 18.14 | 8 | | | 49.94 | 30.14 | 69.73 |
| Asian | | 1 | 241 | | 88.8 | | 84.83 | | 92.77 | | 133 | | 96.24 | | 93.02 | | 99.46 | | 62 | | 85.49 | | 76.81 | | 76.81 | 40 | | | 82.5 | 70.89 | 94.12 |
|  |  | 3 | 214 | | 67.61 | | 61.3 | | 73.91 | | 128 | | 81.93 | | 74.84 | | 89.03 | | 53 | | 52.29 | | 39.16 | | 39.16 | 33 | | | 56.29 | 40.94 | 71.64 |
|  |  | 5 | 113 | | 53.73 | | 45.8 | | 61.66 | | 72 | | 69.01 | | 58.78 | | 79.24 | | 25 | | 40.44 | | 26.34 | | 26.34 | 16 | | | 31.18 | 11.73 | 50.63 |
| Mixed | | 1 | 178 | | 84.83 | | 79.58 | | 90.09 | | 66 | | 93.94 | | 88.23 | | 99.65 | | 52 | | 88.46 | | 79.87 | | 79.87 | 45 | | | 71.11 | 58.05 | 84.18 |
|  |  | 3 | 151 | | 69.92 | | 62.84 | | 77.01 | | 62 | | 85.88 | | 77.34 | | 94.41 | | 46 | | 66.82 | | 52.88 | | 52.88 | 32 | | | 59.14 | 44.73 | 73.54 |
|  |  | 5 | 76 | | 61.17 | | 52.43 | | 69.91 | | 36 | | 76.84 | | 63.01 | | 90.66 | | 20 | | 60.14 | | 44.96 | | 44.96 | 18 | | | 45.54 | 28.33 | 62.75 |
| Other | | 1 | 149 | | 61.75 | | 53.98 | | 69.51 | | 54 | | 85.19 | | 75.81 | | 94.57 | | 47 | | 61.7 | | 48 | | 48 | 28 | | | 57.15 | 39.3 | 74.99 |
|  |  | 3 | 92 | | 46.36 | | 38.2 | | 54.51 | | 46 | | 70.05 | | 57.32 | | 82.77 | | 29 | | 39.79 | | 25.95 | | 25.95 | 16 | | | 41.96 | 23.96 | 59.97 |
|  |  | 5 | 46 | | 38.72 | | 29.46 | | 47.97 | | 25 | | 58.34 | | 42.35 | | 74.32 | | 13 | | 36.17 | | 22.04 | | 22.04 | 7 | | | 33.57 | 14.06 | 53.08 |
| **Cervical cancer** | | | | | | | | | | | | | | | | | | | | | | | | | | | |  |  |  |  |
|  | | | **All ages** | | | | | | | **40-59** | | | | | | | | **60 or over** | | | | | | | | | |  |  |  |  |
| **Ethnicity** | **Follow-up time** | | **Beginning total** | **Net Survival** | | **Lower limit** | | **Upper limit** | | **Beginning total** | | **Net Survival** | | **Lower limit** | | **Upper limit** | | **Beginning total** | | **Net Survival** | | **Lower limit** | | **Upper limit** | | | |  |  |  |  |
| All | 1 | | 2116 | 84.26 | | 82.71 | | 85.82 | | 1285 | | 92.14 | | 90.67 | | 93.61 | | 831 | | 72.09 | | 69.04 | | 75.13 | | | |  |  |  |  |
|  | 3 | | 1783 | 69.49 | | 67.47 | | 71.51 | | 1184 | | 80.81 | | 78.59 | | 83.02 | | 599 | | 51.98 | | 48.47 | | 55.49 | | | |  |  |  |  |
|  | 5 | | 1047 | 64.78 | | 62.51 | | 67.06 | | 743 | | 76.94 | | 74.35 | | 79.52 | | 304 | | 45.98 | | 42.15 | | 49.8 | | | |  |  |  |  |
| White | 1 | | 1871 | 84.13 | | 82.47 | | 85.78 | | 1118 | | 92.76 | | 91.24 | | 94.27 | | 753 | | 71.32 | | 68.09 | | 74.55 | | | |  |  |  |  |
|  | 3 | | 1574 | 69.22 | | 67.07 | | 71.37 | | 1037 | | 81.23 | | 78.89 | | 83.58 | | 537 | | 51.36 | | 47.68 | | 55.05 | | | |  |  |  |  |
|  | 5 | | 942 | 64.57 | | 62.16 | | 66.98 | | 661 | | 77.2 | | 74.44 | | 79.95 | | 281 | | 45.79 | | 41.8 | | 49.77 | | | |  |  |  |  |
| Black | 1 | | 52 | 73.08 | | 61.22 | | 84.94 | | 33 | | 75.76 | | 61.51 | | 90 | | 19 | | 68.43 | | 48.43 | | 88.43 | | | |  |  |  |  |
|  | 3 | | 38 | 63.86 | | 50.45 | | 77.26 | | 25 | | 61.98 | | 45.22 | | 78.75 | | 13 | | 68.44 | | 48.44 | | 88.44 | | | |  |  |  |  |
|  | 5 | | 21 | 63.86 | | 50.45 | | 77.27 | | 15 | | 61.99 | | 45.22 | | 78.75 | | 6 | | 68.45 | | 48.44 | | 88.45 | | | |  |  |  |  |
| Asian | 1 | | 73 | 91.78 | | 85.53 | | 98.04 | | 50 | | 94 | | 87.49 | | 100.52 | | 23 | | 86.96 | | 73.51 | | 100.41 | | | |  |  |  |  |
|  | 3 | | 67 | 74.39 | | 64.19 | | 84.59 | | 47 | | 85.35 | | 75.38 | | 95.33 | | 20 | | 50.18 | | 29.77 | | 70.59 | | | |  |  |  |  |
|  | 5 | | 37 | 64.67 | | 52.22 | | 77.12 | | 29 | | 81.48 | | 69.5 | | 93.45 | | 8 | | 31.36 | | 11.26 | | 51.46 | | | |  |  |  |  |
| Mixed | 1 | | 67 | 91.05 | | 84.26 | | 97.83 | | 44 | | 88.64 | | 79.37 | | 97.9 | | 23 | | 95.65 | | 87.5 | | 103.8 | | | |  |  |  |  |
|  | 3 | | 61 | 80.12 | | 69.89 | | 90.35 | | 39 | | 82.91 | | 71.3 | | 94.52 | | 22 | | 75.56 | | 57.06 | | 94.05 | | | |  |  |  |  |
|  | 5 | | 29 | 75.67 | | 62.97 | | 88.37 | | 20 | | 82.91 | | 71.3 | | 94.52 | | 9 | | 64.76 | | 40.67 | | 88.85 | | | |  |  |  |  |
| Other | 1 | | 53 | 81.13 | | 70.71 | | 91.56 | | 40 | | 90 | | 80.83 | | 99.18 | | 13 | | 53.85 | | 28.19 | | 79.51 | | | |  |  |  |  |
|  | 3 | | 43 | 62.52 | | 48.05 | | 77 | | 36 | | 75.43 | | 60.5 | | 90.36 | | 7 | | 18.46 | | -3.5 | | 40.46 | | | |  |  |  |  |
|  | 5 | | 18 | 59.05 | | 43.94 | | 74.16 | | 18 | | 71.24 | | 55.15 | | 87.33 | | 0 | | 0 | | 0 | | 0 | | | |  |  |  |  |

*Net survival estimates represent the probability of surviving cancer in the absence of other causes of death. Estimates are shown at 1, 3, and 5 years after diagnosis. “Beginning total” refers to the number of individuals at risk at the start of each follow‑up interval. The estimates reported here were not standardised, lower and upper limits represent the corresponding 95% confidence intervals. The estimates for ovarian and cervical cancers were limited to fewer age bands due to small number of cases by ethnicity.*

**S6. Ethnic differences in the proportion of current smokers at diagnosis by cancer type**

Percent of current smoker

|  | All ethnicities | White | Black | Asian | Mixed | Other |
| --- | --- | --- | --- | --- | --- | --- |
| Breast | 14.07 | 14.38 | 11.76 | 5.74 | 16.24 | 13.09 |
| Lung | 38.39 | 38.25 | 45.67 | 25.48 | 42.89 | 44.85 |
| Prostate | 12.18 | 11.8 | 18.42 | 11.47 | 13.77 | 14.58 |
| CRC | 13.45 | 13.39 | 14.51 | 11.85 | 14.73 | 15.03 |
| OG | 21.36 | 21.34 | 22.61 | 17.79 | 23.13 | 22.22 |
| Cervix | 25.8 | 26.67 | 15.38 | 6.85 | 34.33 | 20.37 |
| Myeloma | 11.91 | 11.66 | 15.58 | 7.73 | 15.64 | 12.5 |
| Ovary | 13.05 | 13.42 | 10.78 | 2.48 | 14.53 | 14.29 |
| All site | 18.6 | 18.7 | 18.81 | 10.94 | 20.23 | 20.23 |

*Percentages reflect the proportion of individuals who were current smokers at the time of diagnosis, stratified by cancer type and ethnicity. Error bars = 95% confidence intervals; OG =Oesophagogastric; error bars=95% confidence interval*

**S7. Ethnic differences in the proportion of ex- and never-smokers at diagnosis by cancer type**

|  | All ethnicities | White | Black | Asian | Mixed | Other |
| --- | --- | --- | --- | --- | --- | --- |
| Breast | 62.1 | 61.7 | 65.3 | 70 | 64.9 | 59.2 |
| Lung | 45.9 | 46.1 | 38.8 | 54.1 | 44.7 | 37.4 |
| Prostate | 66.2 | 66.5 | 60.3 | 67.8 | 69.3 | 56.8 |
| CRC | 64.1 | 64.2 | 61.5 | 66.1 | 67 | 56.5 |
| OG | 58.9 | 59.3 | 55.8 | 59.7 | 60.7 | 47.7 |
| Cervix | 49.5 | 48.2 | 67.3 | 71.2 | 52.2 | 44.4 |
| Myeloma | 66 | 66.2 | 62.3 | 67.2 | 69.2 | 61.5 |
| Ovary | 62.5 | 62.4 | 63.7 | 65.3 | 67 | 55.8 |
| All site | 60.2 | 60.1 | 59.6 | 66.1 | 62.9 | 53.2 |

*Percentages reflect the proportion of individuals who were ex-smokers or never-smokes at the time of diagnosis, stratified by cancer type and ethnicity. Error bars = 95% confidence intervals; OG =Oesophagogastric; error bars=95% confidence interval*
